# Allele-specific expression of *ATXN3* in blood samples of Machado-Joseph disease expansion carriers

**DOI:** 10.64898/2026.01.26.26344891

**Authors:** Ana Rosa Vieira Melo, Sandra Martins, Sara Pavão, Luís Teves, Ana F. Ferreira, Ahmed M. Sidky, Jorge Sequeiros, Darren G. Monckton, Manuela Lima, Mafalda Raposo

**Author notes:** Corresponding author: Mafalda Raposo Instituto de Investigação e Inovação em Saúde (i3S) Rua Alfredo Allen, 208 4200-135 Porto, Portugal. Both authors contributed equally to this work (Ana Rosa Vieira Melo, Sandra Martins).

## Abstract

Although the CAG repeat expansion in the *ATXN3* gene was identified over 30 years ago as the cause of Machado-Joseph disease (MJD), the disorder remains untreatable. Notably, MJD is the most prevalent hereditary spinocerebellar ataxia worldwide and is particularly frequent in the Azores Islands (Portugal). This results from two independent founder effects, with two major ancestral lineages – “Joseph” and “Machado” - segregating in Azorean families. Although MJD pathogenesis is mainly driven by the (CAG)_n_ expansion, regulatory *cis-*elements may modulate *ATXN3* expression, thereby influencing phenotypic variation. Here, we investigated allele-specific expression (ASE) of the *ATXN3* gene and examined whether distinct *ATXN3* lineages modulate the differential expression of non-expanded and expanded alleles, as well as its association with age at onset. We quantified *ATXN3* ASE in 38 cDNA samples from the blood of Azorean MJD expansion carriers. Notably, all 28 carriers of the Joseph lineage demonstrated higher relative expression of the expanded allele, whereas eight of the 10 Machado lineage carriers exhibited the opposite trend (Mann-Whitney U test, *p* < 0.0001). By incorporating genetic and clinical data from an additional 76 Azorean MJD patients we found that, based on the expanded allele size alone, Joseph carriers would be expected to develop symptoms later than Machado carriers. Both lineages, however, report similar ages at onset. suggesting a counterbalance effect of *ATXN3* ASE: higher expression of the expanded allele in Joseph carriers may shift individuals toward earlier onset, while higher expression of the non-expanded allele in Machado carriers, may contribute to delayed onset. Our findings show that *ATXN3* exhibits haplotype-dependent allelic imbalance. This imbalance may be tissue-specific, underscoring the need for future studies using brain samples. Furthermore, it will be important to determine whether the observed association with age at onset is driven primarily by increased levels of the expanded protein, leading to enhanced protein toxicity, or by toxic effects at the RNA level.

## 1. INTRODUCTION

Machado-Joseph disease/spinocerebellar ataxia type 3 (MJD/SCA3) is the most prevalent spinocerebellar ataxia worldwide and remains with no effective treatment. An expanded coding (CAG)_n_ tract in the *ATXN3* gene above 60 repeats is translated into an elongated glutamine tract in the ubiquitous ataxin-3 protein, triggering the disease, mainly by a toxic gain-of-function (reviewed in Bettencourt & Lima, 2011). The clinical phenotype of MJD includes a progressive cerebellar ataxia, dysarthria and oculomotor abnormalities, accompanied by involvement of other neurological systems, such as pyramidal and extrapyramidal signs, peripheral neuropathy, spasticity, and, in some cases, parkinsonism or dystonia (Coutinho, 1992). Age at onset (∼40 years) and clinical presentation are highly variable, partly reflecting differences in CAG repeat length and genetic background (e.g., Elter et al., 2025; Raposo et al., 2015). Ataxia severity and progression as well as non-ataxic manifestations are commonly assessed using the scale for the assessment and rating of ataxia (SARA) and the inventory of non-ataxia signs (INAS), respectively (Jacobi et al., 2013; Schmitz-Hübsch et al., 2006).

Mutant ataxin-3 is involved in the dysregulation of several molecular and cellular mechanisms, such as transcription, translation, and DNA repair (reviewed in Da Silva et al., 2019). Although the toxic effects of the expanded ataxin-3 protein are relatively well established (reviewed in Evers et al., 2014), the contribution of RNA toxicity in MJD pathogenesis remains largely unexplored. Studies have investigated the potential involvement of alternative splicing, bidirectional transcription, RNA interference pathway, as well as repeat-associated non-ATG-initiated translation in MJD and in the pathology of other polyglutamine expansion disorders (reviewed in (Evers et al., 2013; Nalavade et al., 2013; Raposo et al., 2024; Rohilla & Gagnon, 2017). Allele-specific expression (ASE), a mechanism of gene regulation in which one allele at a specific locus is preferentially expressed over the other (*e.g.*, St. Pierre et al., 2022) has not been studied in MJD. An allele-specific approach is a powerful tool to study imbalance caused by *cis*-regulatory genetic variation and epigenetic alterations, such as imprinting (Castel et al., 2020). Although MJD pathogenesis is mainly driven by the (CAG)_n_ expansion, regulatory cis elements may affect *ATXN3* expression, thereby modulating phenotypic variation.

Suppression of *ATXN3* expression has been successfully performed *in vivo* and *in vitro* models of MJD (*e.g.*, Santos et al., 2023; Wu et al., 2015), justifying the recruitment of MJD patients for a phase I/II trial to assess the safety and tolerability of a new ASO-based drug developed by Vico Therapeutics, currently ongoing (NCT05822908). The use of expanded ataxin-3 protein levels as a target engagement biomarker in plasma has been demonstrated (Hübener-Schmid et al., 2021); however, misfolded and aggregated proteins can interfere with accurate quantification (Liu et al., 2013). Additionally, this approach does not account for the assessment of non-coding RNA species, such as lncRNAs (such as the ones recently described by us for *ATXN3;* Raposo et al., 2024), which may also play a role in disease pathogenesis. So far, quantification of ataxin-3 RNA levels has been limited by the inability to measure *ATXN3* expression in an allele-specific manner, a constraint that current NGS technology has not yet overcome. Single-nucleotide variants (SNVs) within coding and untranslated regions (UTRs) of *ATXN3*, for which patients are frequently heterozygous (Goto et al., 1997; Maciel et al., 1999; Melo et al., 2023; Ogun et al., 2015), offer an opportunity to discriminate between non-expanded and expanded *ATXN3* alleles. In patients, this heterozygosity depends on the haplotype background (MJD lineage) inherited in the family, as well as on the allele frequency of SNVs observed in the respective population. In MJD, two major SNV-based haplotypes have been shown to segregate with expanded *ATXN3* alleles (Martins & Sequeiros, 2018). The Joseph lineage is globally widespread (including mainland Portugal and the Azorean Island of Flores), whereas the Machado lineage is only frequently observed in mainland Portugal and in the Azorean Island of Sao Miguel, being rare in other populations (Gaspar et al., 1996; Martins et al., 2007, 2012; Sharony et al., 2019).

Recently, non-expanded and expanded *ATXN3* mRNA levels were determined in MJD cell lines using droplet digital PCR (ddPCR; Joachimiak et al., 2023). Lower expression levels of the expanded *ATXN3* allele compared to the wild-type allele were observed in one patient-derived fibroblast cell line that was subsequently reprogrammed into induced pluripotent stem cells (iPSCs) and differentiated into neural stem cells and neurons (Joachimiak et al., 2023). However, allele-specific expression analysis in human tissues from MJD expansion carriers has not yet been performed.

In this study, we developed a novel quantitative PCR (qPCR)-based protocol to determine the expression ratio of expanded to non-expanded *ATXN3* alleles in blood samples from MJD subjects by distinguishing allelic mRNA species based on the heterozygous status of SNVs in the *ATXN3* gene. We further tested if the differential expression of normal and expanded *ATXN3 is* associated with distinct *ATXN3* lineages, and whether these modulate the age at onset of the disease.

## 2. SAMPLES AND METHODS

### 2.1. Subjects and samples

Whole blood-derived DNA and cDNA samples from a total of 224 carriers of the *ATXN3* expansion of Portuguese ancestry were used in this study; 116 MJD expansion carriers were from the Azorean cohort (Lima et al., 2023), whereas the remaining 108 were from the mainland cohort. Ninety cDNA samples of Azorean MJD expansion carriers were used in the protocol development; DNA samples from Azorean and mainland MJD cohorts were used to determine genotype frequencies of SNVs in the *ATXN3* gene. cDNA was previously obtained as described in Raposo and colleagues (Raposo et al., 2015). DNA was obtained as previously described by Melo and colleagues (Melo et al., 2022) and Costa and colleagues (Costa et al., 2019) from samples of Portuguese Azorean and mainland MJD subjects, respectively. For each Azorean subject, the age at blood collection, the number of CAG repeats in the *ATXN3* and the age at onset were available. Age at visit (= age) was determined by subtracting the year of birth from the year of blood collection. Age at onset was defined as the age at which the patient, a close relative, or a caregiver first reported gait disturbance. The number of CAG repeats in *ATXN3* was determined as described by Sidky and colleagues (Sidky et al., 2024).

The study was approved by local ethics committees (Universidade dos Açores and Instituto de Investigação e Inovação em Saúde da Universidade do Porto) and all subjects provided written informed consent.

### 2.2. Protocol design

The quantitative PCR (qPCR)-based protocol to determine the relative expression ratio of expanded to non-expanded *ATXN3* alleles comprised two main steps: (1) – identification of heterozygotes for the SNV of interest and quantification of allele-specific transcripts by qPCR; and (2) – determination of the SNV-(CAG)_n_ phasing through electrophoresis followed by Sanger sequencing of DNA fragments from both larger (expanded allele) and shorter (non-expanded allele) fragments. Before steps 1 and 2, three key preparatory steps were carried out to ensure control over critical methodological factors, such as cDNA input and TaqMan assay performance. The protocol was developed and optimized using the SNV rs1048755 (GRCh38.p13, ENST00000644486.2:c.634G>A), which was randomly selected amongst the four SNVs with minor allele frequency >1%, as described in Ensembl (Dyer et al., 2025). An illustrative scheme of the protocol is shown in Figure 1. A comprehensive description of the protocol development is available in the supplementary data.

**Figure 1.**
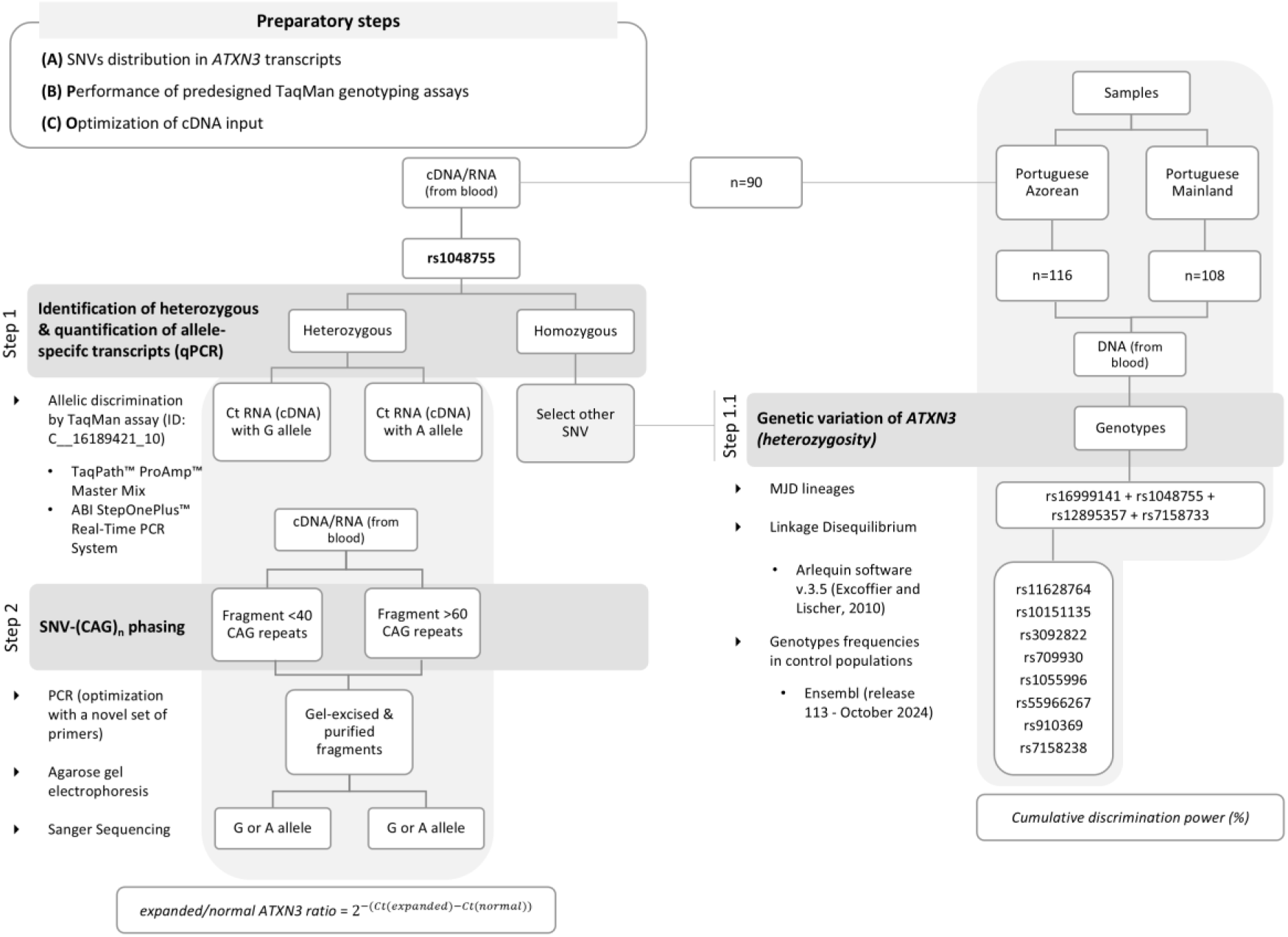
Workflow illustrating all steps involved in the development of a protocol for determining allele-specific expression (ASE) of *ATXN3*. The protocol comprises two main steps: (1) identification of heterozygotes for rs1048755 and quantification of allele-specific transcripts by qPCR, and (2) determination of SNV–(CAG)n phasing via electrophoresis and Sanger sequencing of DNA fragments corresponding to expanded and non-expanded alleles. Prior to this, three preparatory steps were performed to investigate critical methodological variables. To identify heterozygotes for additional *ATXN3* SNVs, an extra step (1.1) was introduced, involving analyses of 11 SNVs. RNA or DNA samples from the Portuguese Azorean and mainland MJD cohorts were used.

### 2.3. Identification of MJD lineages

MJD lineages were assessed using the SNV-based haplotype (rs16999141-rs1048755-rs12895357-rs7158733) in *cis* with expanded CAG alleles. To infer haplotypes, we genotyped family members (whenever available), which allowed us to assess allelic phases of the four SNVs and the (CAG)n by segregation.

The complete haplotype was successfully assessed in 16 out of the 26 families analyzed; heterozygosity did not allow us to infer allelic phases in six families for rs7158733, three families for rs16999141, six families for rs1048755, and eight families for rs12895357. In one Joseph family and two Machado families, we found one of the four SNVs did not share the allele commonly segregating with the expansion. Since the remaining haplotype (with three SNVs) was successfully inferred in these families, we included them in the analysis.

### 2.4. *In silico* analysis of the SNVs rs16999141, rs1048755, rs12895357, rs7158733

Information on the predicted functional consequences of SNVs was obtained using the Ensembl Variant Effect Predictor (VEP) web interface (McLaren et al., 2016) with genome assembly GRCh38.p14 and the Ensembl/GENCODE transcript database selected as standard parameters. Potential effects of the variants on RNA splicing were evaluated using Human Splicing Finder (HSF) system (Desmet et al., 2009) a tool designed to identify core and auxiliary splicing signals, including acceptor and donor splice sites, branch points and exonic splicing enhancers (ESEs) and silencers (ESSs). All *in silico* analyses were performed at the transcript level. However, the evaluation was restricted to *ATXN3* transcripts known to be frequently expressed in blood, as previously described by Raposo and colleagues (Raposo et al., 2024).

### 2.5. Statistical analysis

Normality of continuous variables was assessed using the Shapiro-Wilk test. Nonparametric tests were employed for variables that were not normally distributed or whenever the sample size was small. The relative expression ratio of expanded to non-expanded *ATXN3* alleles across individuals was compared by the one-sample Wilcoxon test. Differences between sex or island of origin and the expression ratio were determined by the Mann-Whitney U or by the Kruskal-Wallis test, respectively. To explore the direction and strength of the relationship between expression ratio and demographic (age), genetic data (number of CAG repeats in the *ATXN3*), and clinic (AO), Spearman correlation coefficients (rho) were calculated. Fisher’s exact test was used to compare the proportion of subjects by expression ratio trends and MJD lineage. *ATXN3* expression ratio was compared between MJD lineages using a Mann-Whitney U test. A multiple linear regression was conducted to examine whether sex, age, AO, non-expanded allele, expanded allele, and MJD lineages predicted *ATXN3* ASE. An ANCOVA, using the expanded allele as a covariate, was performed to calculate and compare estimated age at onset between MJD lineages. A linear regression model was used to quantify the extent to which the number of CAG repeats in the expanded allele explains age at onset variance and to generate corresponding predictions of age at onset. Differences between the CAG repeat length in the non-expanded and expanded alleles, as well as the reported and predicted ages at onset and MJD lineages, were determined by the Mann-Whitney U test. The relative frequency distributions of these variables were also compared using a Fisher’s exact test. Statistical analyses were performed in IBM SPSS Statistics for Windows version 25.0 (IBM Corp. Released 2017) and GraphPad Prism 8.0.1. The significance level of all tests was set to 5%. To control Type I errors, *post hoc* analyses using Dunn’s multiple-comparison test were performed.

## 3. RESULTS

Our novel protocol was successfully implemented for allele-specific expression analysis of *ATXN3* in blood samples of MJD expansion carriers. SNV rs1048755 was heterozygous in 44 out of the 90 cDNA samples used (frequency of 0.5). For a subset of 24 samples, we determined the rs1048755-(CAG)_n_ phase, enabling us to successfully calculate the relative expression ratio of expanded to non-expanded *ATXN3* alleles. By integrating rs12895357-(CAG)_n_ phasing information from a previous study (Ahmed M Sidky et al., 2024) with rs1048755-(CAG)_n_ phasing from our results, we discriminated and determined the relative expression ratio of expanded to non-expanded *ATXN3* alleles in 14 additional samples. In total, allele-specific *ATXN3* levels were available for 38 samples. The expanded allele was expressed at levels 1.2 times higher than the non-expanded allele (95% CI 1.107 to 1.252; one sample Wilcoxon test, *P* < 0.001; Figure 2a). Notably, carriers of the MJD expansion from Flores and Terceira islands showed, on average, higher expression of the expanded allele relative to the non-expanded allele (ratio > 1). In contrast, carriers exhibiting on average higher expression of the non-expanded allele (ratio < 1) were from São Miguel and Graciosa islands (Figure 2c). This geographic pattern supports the hypothesis that island-specific haplotypes, first reported nearly three decades ago (Gaspar et al., 1996; Martins et al., 2007; Martins & Sequeiros, 2018), contribute to variation in allele-specific expression (ASE) among islands. Specifically, patients from Flores and Terceira typically carry the Joseph MJD lineage, whereas patients from São Miguel and Graciosa predominantly exhibit the Machado background. No significant correlations were found between *ATXN3* ASE and sex, age at evaluation, CAG repeat size in the expanded allele, or age at onset (Figure 2d). Interestingly, the CAG repeat size in the non-expanded allele showed a significant negative association with allelic expression *i.e.*, individuals with longer non-expanded alleles exhibited lower ASE ratios (Spearman rank correlation, p=0.021). This suggested that longer non-expanded alleles might be expressed at higher levels than the expanded allele.

**Figure 2.**
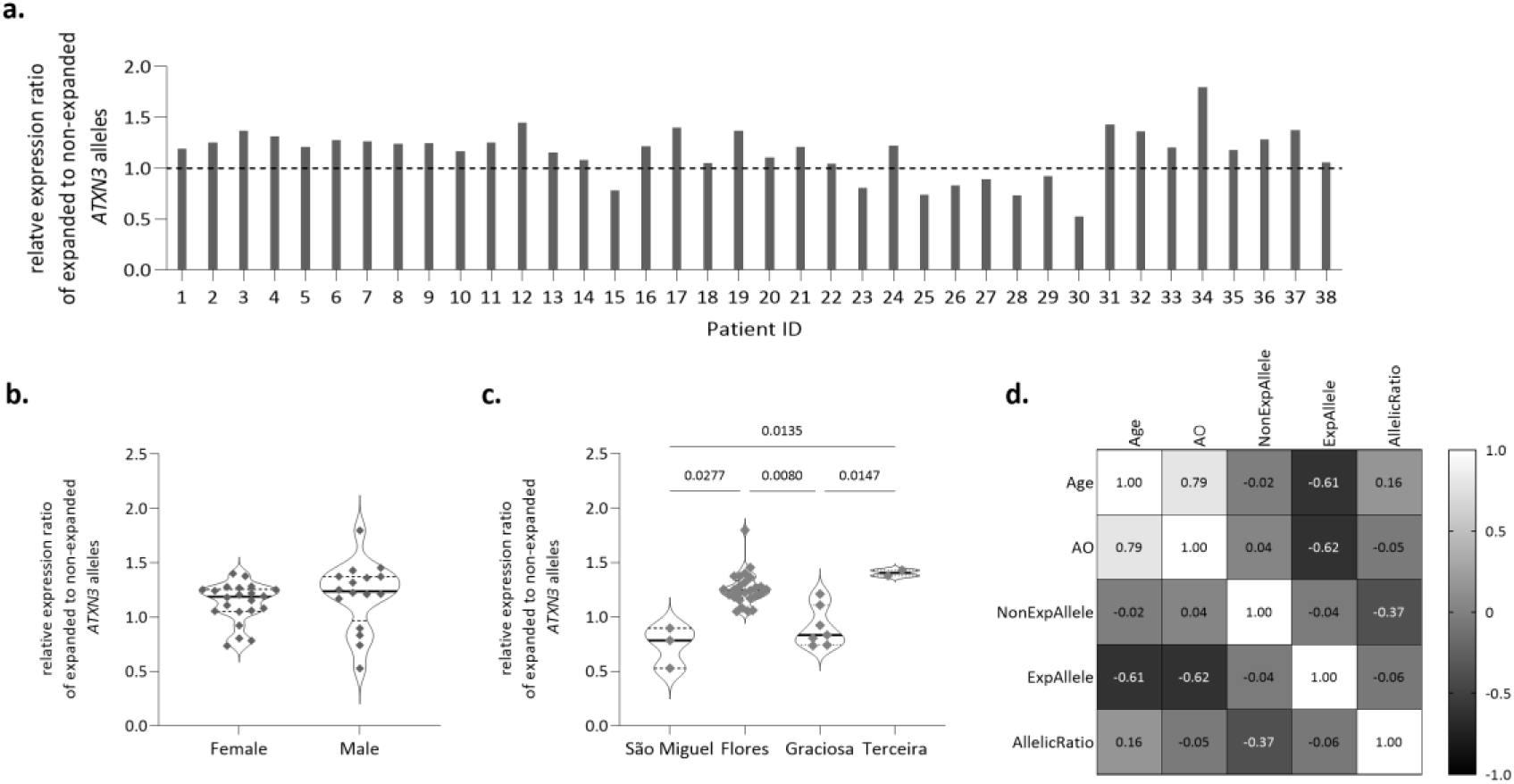
*ATXN3* allele-specific expression on MJD. (a) The relative expression ratio of expanded to non-expanded *ATXN3* alleles in 38 MJD blood samples. The dashed line represents a ratio of 1, indicating equal expression levels of the non-expanded and expanded alleles. Comparison of *ATXN3* ASE between (b) sex and (c) island of origin. (d) Spearman rank correlation matrix of age at evaluation (Age), age at onset (AO), the number of CAG repeats in non-expanded (NonExpAllele) and expanded (ExpAllele) alleles with *ATXN3* ASE (AllelicRatio).

### Differential imbalance of ATXN3 allele-specific expression in the Joseph and the Machado lineages

The MJD lineages, Joseph (Flores and Terceira Islands) and Machado (São Miguel and Graciosa Islands), have been characterized by distinct haplotypes in *cis* with the expanded allele, as determined by analysis of six SNVs within the *ATXN3* gene, including rs1048755 and rs12895357 (Gaspar et al., 1996; Martins et al., 2007; Martins & Sequeiros, 2018). Based on this haplotype, we assigned each MJD carrier to its respective lineage. In close agreement with their geographic distribution, all 28 carriers of the Joseph lineage showed a higher level of expression of the expanded alleles, whilst eight of the 10 Machado lineage carriers displayed the opposite trend, suggesting a possible haplotype-specific effect (Fisher’s exact test, *p* < 0.0001; Figure 3a). Indeed, there was a highly statistically significant difference in median *ATXN3* ASE levels between the two MJD haplotypes (Mann-Whitney U test, *p* < 0.0001; Figure 3b).

**Figure 3.**
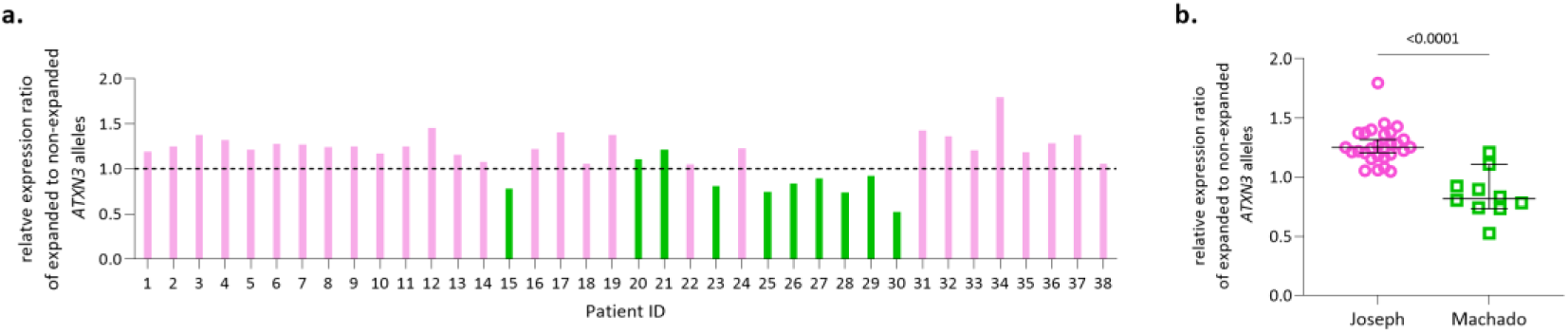
Expanded allele haplotype-dependent *ATXN3* allele-specific expression in MJD. The relative expression ratio of expanded to non-expanded *ATXN3* alleles in 38 MJD blood samples is shown (a). Graphic bars in pink represent MJD subjects from the Joseph lineage, while bars in green represent MJD subjects from the Machado lineage. The dashed line represents a ratio of 1, indicating equal expression levels of the non-expanded and expanded alleles. (b) Comparison of *ATXN3* ASE between MJD lineages – Joseph or Machado. Graphic bars are shown as median ± 95% CI (confidence interval).

To understand the opposite tendency in the relative expression ratio of expanded to non - expanded *ATXN3* alleles between MJD lineages, we investigated demographic, genetic, and clinical data in both groups (Supplementary Figure 1). Sex, age at evaluation, CAG repeat size in the expanded allele, and age at onset – reported or adjusted (controlled for the CAG expansion length) - were similar in MJD subjects from the Joseph and Machado lineages (Supplementary Figure 1). The CAG repeat size of the non-expanded allele was significantly larger in carriers of the Machado lineage than in those of the Joseph lineage (Mann-Whitney test, p = 0.006; Supplementary Figure 1). A multiple linear regression was conducted to examine if independent variables predicted *ATXN3* ASE (*ATXN3* ratio ∼ intercept + sex + age + AO + non-expanded allele + expanded allele + MJD lineages). The model was statistically significant, F(6,24)=12.1, P<0.0001, and explained 75% of the variance in *ATXN3* allelic expression (r^2^=0.751, adjusted r^2^=0.689). Only MJD lineages significantly predicted *ATXN3* ASE (B=-0.489, SE=0.074, t(24)=6.577, P<0.001), with Machado patients showing a lower ratio of allelic expression levels than Joseph patients.

Findings on the association between MJD lineages and *ATXN3* ASE prompted further investigation into the potential contribution of MJD haplotypes to the disease phenotype. To explore this relationship, genetic and clinical data from an additional 76 Azorean carriers of MJD expansions were included in the analysis. The average of the CAG repeat length in both non-expanded and expanded alleles, as well as the reported and CAG-predicted ages at onset, were similar between carriers of the Joseph and Machado lineages (Figure 4a-d). However, the relative frequency distributions of the CAG repeat length in expanded allele and the CAG-predicted age at onset, differed significantly between the two MJD lineages (Chi-squared test, p=0.0003 and p<0.0001, respectively; Figure 4b1, d1). Machado subjects presented a higher frequency of longer non-expanded and expanded alleles, compared to carriers of the Joseph lineage (Figure 4a1, b1). Therefore, as expected, an earlier predicted age at onset was more frequent among carriers of the Machado lineage than among the Joseph lineage (Figure 4d1). However, the reported age at onset remained similar between carriers of both lineages (Figure 4c1). Among carriers of the Joseph lineage, 48% of the variance in age at onset was explained by the CAG repeat length in the expanded allele, whereas in carriers of the Machado lineage, this proportion increased to 59% (Figure 4e). Despite these differences, the slopes of the association between CAG repeat length in the expanded allele and age at onset was similar for both MJD lineages.

**Figure 4.**
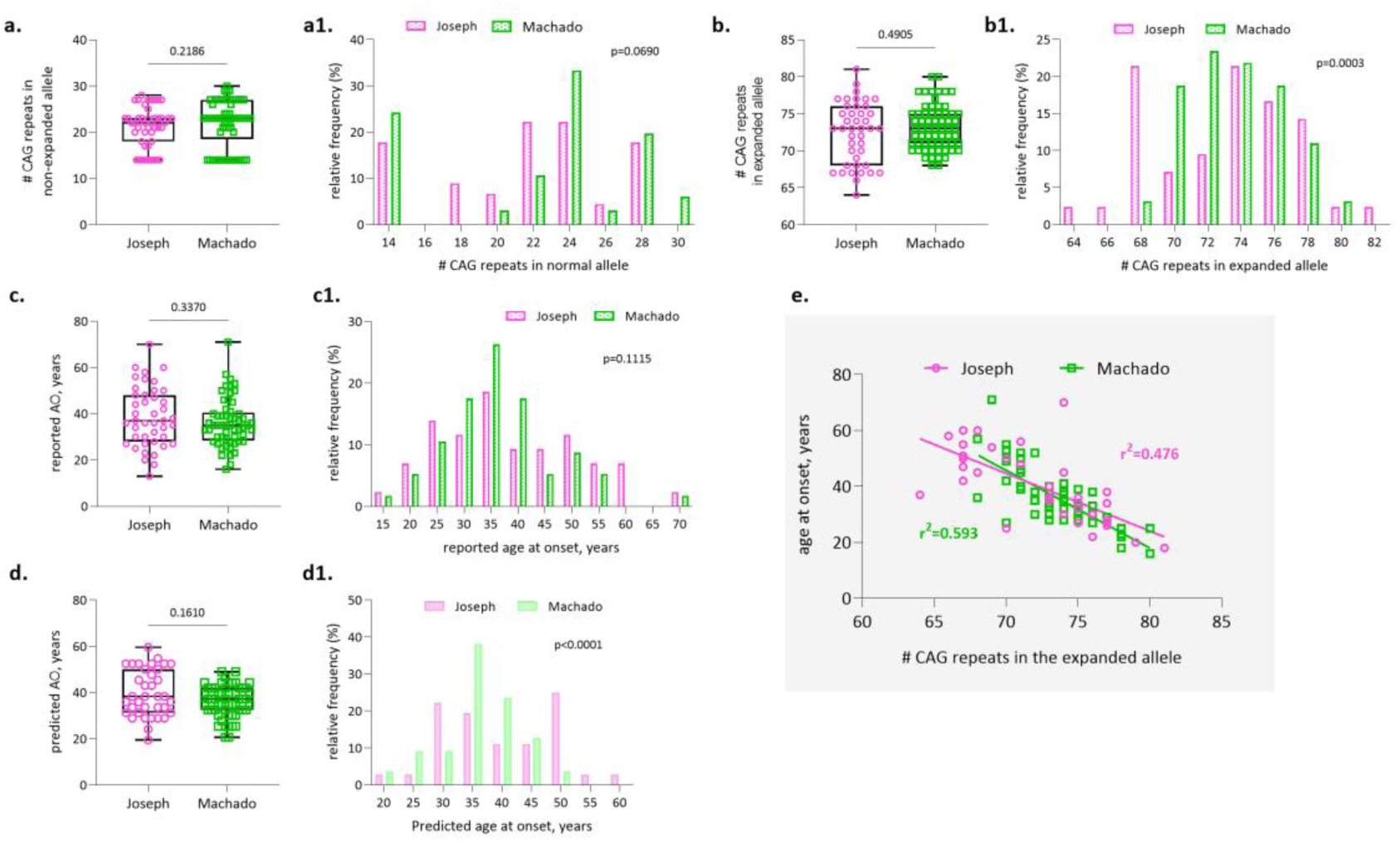
Genetic and clinical characterization of the Azorean MJD cohort (n = 114). Data for the CAG repeat length in the non-expanded and expanded alleles, as well as the reported and predicted ages at onset are presented as box plots (a-d) or as histograms showing relative frequencies (a1-d1). For histogram visualization, data were grouped into discrete intervals (bins) to facilitate clearer representation and interpretation. Linear regression analyses were performed separately for each MJD lineage to evaluate the association between CAG repeat length in the expanded allele and age at onset (e).

### Pathogenicity prediction suggests that rs16999141 and rs7158733 may alter protein function or RNA splicing

Predictions of the potential impact of SNVs rs16999141, rs1048755, rs12895357, and rs7158733 were performed to assess (i) their effects on the structure and function of the respective protein isoform, and (ii) their influence on splicing signals. The analysis was restricted to *ATXN3* transcripts frequently expressed in blood, including two protein-coding transcripts (*ATXN3-251, ATXN3-214*), two transcripts targeted for NMD (*ATXN3-202, ATXN3-254*), one transcript with a protein CDS not defined (*ATXN3-249*), and three retained-introns (*ATXN3-210, ATXN3*–205, *ATXN3*–248) transcripts (Raposo et al., 2024). SNVs rs16999141, rs1048755, rs12895357, and rs7158733 are located in distinct regions of the *ATXN3* gene, depending on the transcript analyzed (Figure 5). The four SNVs have no predicted impact on the *ATXN3-249, ATXN3-210, ATXN3*–205, and *ATXN3*–248 transcripts, as their sequences do not include the genomic regions in which these SNVs are located (Figure 5). Therefore, only *ATXN3-251, ATXN3-214, ATXN3-202,* and *ATXN3-254* were considered for further analysis.

**Figure 5.**
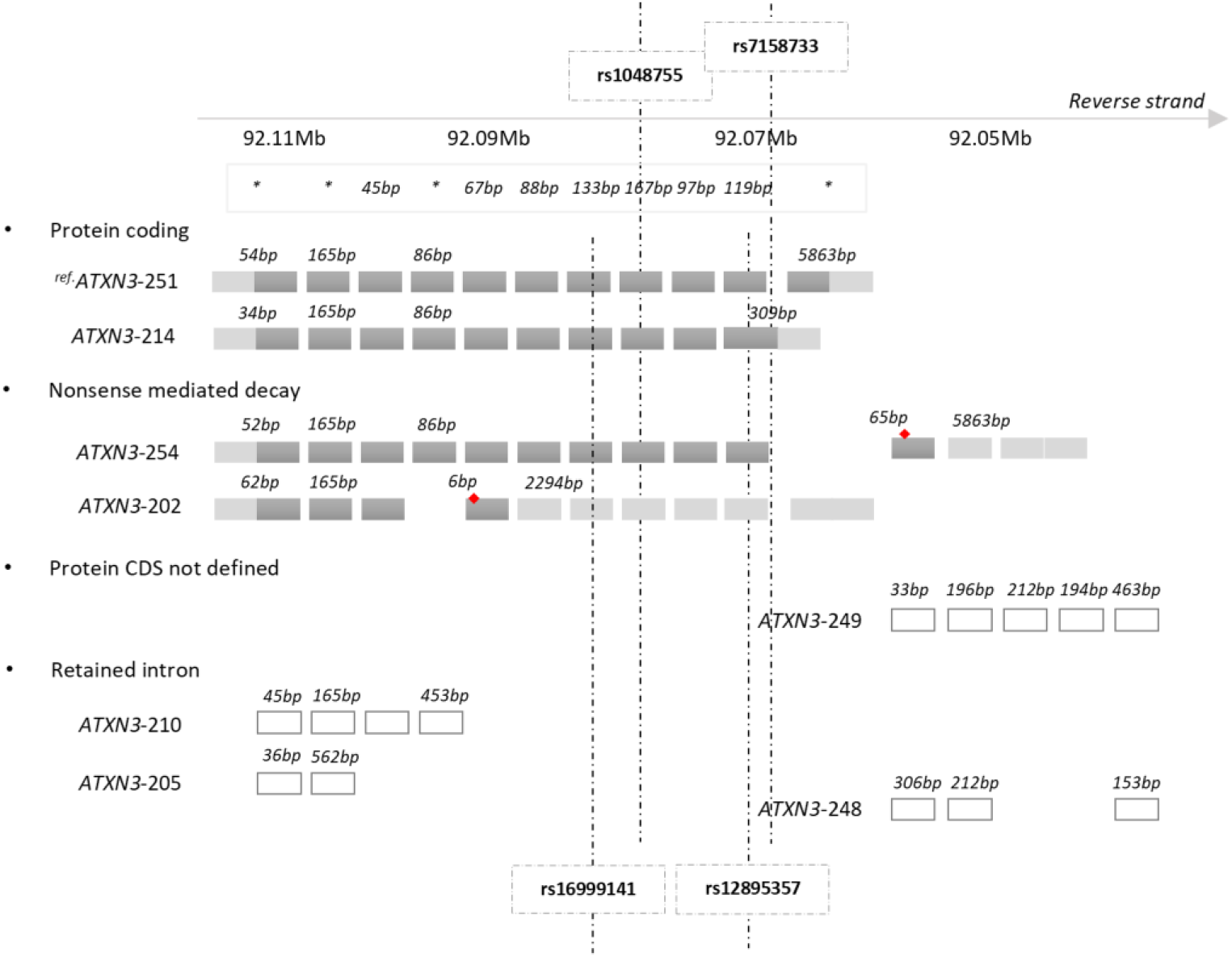
Schematic representation of the localization of the four SNVs rs16999141, rs1048755, rs12895357, and rs7158733 in the eight annotated *ATXN3* transcripts frequently expressed in blood samples from MJD expansion carriers and controls (adapted from (Raposo et al., 2024)). *ATXN3* transcripts are grouped according to Ensembl biotype - protein coding, nonsense mediated decay, protein coding sequence (CDS) not defined, and retained intron. Ref indicates Ensembl *ATXN3* reference transcript ENST00000644486.2|ATXN3-251 (MANE Select v0.95). Red diamonds denote premature termination codons. Grey boxes represent coding exons, light grey boxes untranslated regions (UTRs), and white boxes non-coding exons. Exon, intron, and UTR sizes are not drawn to scale. *Exon size varies across *ATXN3* transcripts.

Considering the predicted consequences of the four SNVs on the structure and function of the corresponding protein isoforms, only rs7158733 is classified as having a high impact, as it introduces a premature stop codon. Notably, this effect is specific to the *ATXN3-214* transcript (Table 1). A potential alteration in splicing was predicted for rs7158733, driven by the activation of a cryptic acceptor splice site in the *ATXN3-251, ATXN3-214,* and *ATXN3-254* transcripts. In addition, rs16999141 was predicted to cause a substantial change in splicing regulatory elements, reflected by a marked reduction in the ESE/ESS motifs ratio (−10) in the same three transcripts (Table 1).

**Table 1.**
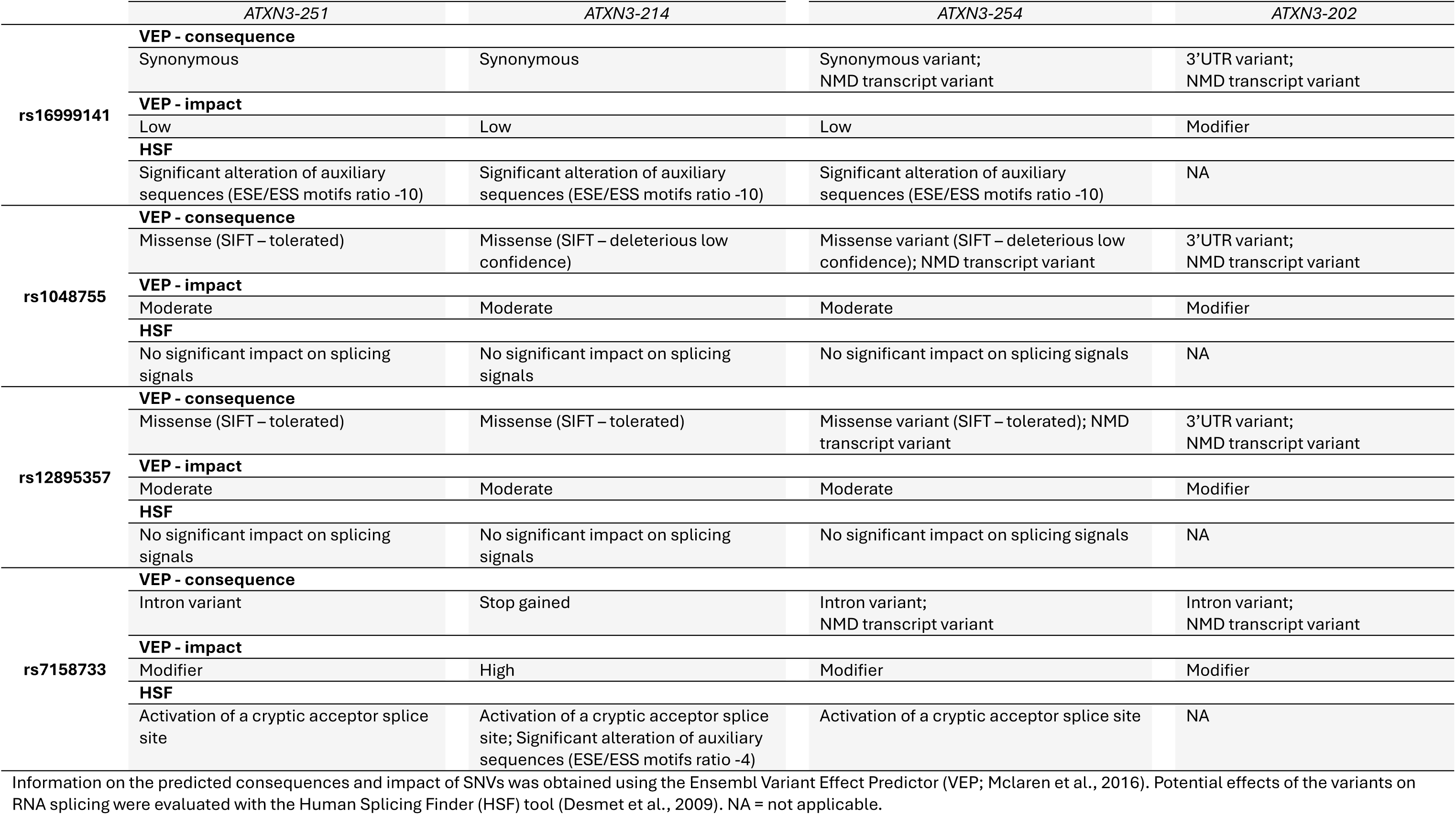
***In silico* analysis of the effects of the SNVs rs16999141, rs1048755, rs12895357 and rs7158733 on the structure and function of the respective protein isoform as well as on splicing signals.**

## 4. DISCUSSION

In this study, we aimed to document allele-specific expression (ASE) of the *ATXN3* gene in blood samples from individuals with Machado–Joseph disease (MJD), and to examine whether differences in ASE are associated with the two major Machado–Joseph genetic lineages and with variability in age at disease onset. Based on the heterozygous status of single nucleotide variants (SNVs) at the *ATXN3* gene, we developed a protocol allowing the effective discrimination of non-expanded and expanded *ATXN3* alleles and the quantification of the relative transcript levels in blood samples of MJD expansion carriers. We also define a set of five assayable SNVs, with a combined heterozygosity of ∼59% in the Portuguese MJD cohort, to maximize the number of MJD expansion carriers that can be included in subsequent analyses of *ATXN3* allele-specific expression. If additional cohorts from other MJD populations are analyzed, this protocol can be adapted by selecting the most informative SNVs based on population-specific allele frequencies and the distribution of MJD lineages in the population under study. Overall, *ATXN3* allele-specific expression is unbalanced, with the expanded allele being, on average, relatively more highly expressed than the non-expanded allele. Notably, our results show that the *ATXN3* expression ratio differs according to MJD lineage: in subjects carrying the Joseph lineage, the expanded *ATXN3* allele is more highly expressed than its non-expanded counterpart, whereas in subjects carrying the Machado lineage, the opposite trend was observed.

Few studies have explored the abundance of *ATXN3* at the RNA level (Joachimiak et al., 2023; Nishiyama et al., 1996). The first study, restricted to the analysis of total *ATXN3* levels by *in situ* hybridization, confirmed ubiquitous expression of *ATXN3,* by using different brain tissues of MJD patients and controls, as well as other tissues, such as heart, liver, spleen, and others (Nishiyama et al., 1996). Recently, allele-specific expression of *ATXN3* was analyzed in cell models of MJD using a SNP-based quantitative droplet digital PCR protocol (Joachimiak et al., 2023). Compared to the non-expanded allele, the expanded allele showed lower expression levels in fibroblasts, iPSCs reprogrammed from fibroblasts, and neural stem cells (NSCs) and neurons differentiated from iPSCs. The fibroblasts were derived from a single patient with the Joseph lineage (Joachimiak et al., 2023); however, we could not confirm this finding in our study, since all carriers of the Joseph lineage we analyzed showed a relatively higher expression of the expanded allele.

Although mRNA levels do not always predict protein abundance, due to post-transcriptional or post-translational regulatory mechanisms (*e.g.,* Vogel & Marcotte, 2012), our findings align with the trend observed at the protein level. The ratio of the expanded to non-expanded ataxin-3 protein levels was quantified in a small set of peripheral blood mononuclear cells (PBMC) samples from 10 MJD patients using a highly specific fluorophore-labeled ataxin-3 antibody (Breuer et al., 2022). The expanded protein was found at higher levels than its non-expanded counterpart, in all samples (Breuer et al., 2022). Breuer and colleagues also assessed correlations between ataxin-3 protein ratio and age, the number of CAGs in the expanded allele, age at onset and SARA score. A significant negative correlation with age at onset was observed: patients with a higher relative abundance of expanded ataxin-3 in PBMCs tended to have earlier disease onset (Breuer et al., 2022). In our cohort (n=38 carriers of MJD expansion), we did not find a significant correlation between transcript levels and age at onset. Although age at onset is the most used phenotypic variable in genotype-phenotype correlations (*e.g.*, Akçimen et al., 2020; Raposo et al., 2021; Weber et al., 2014), it is often self-reported and subject to recall bias. Additionally, age at onset is not solely determined by the size of the CAG repeats, an association varying across cohorts (*e.g.*,de Mattos et al., 2019). Future studies incorporating more precise phenotypic variables, namely ataxia severity assessed by clinical scales, such as SARA, will enhance the accuracy and reliability of phenotype-genotype correlations. Additionally, it will be important to consider non-ataxic features as possible initial symptoms of disease onset. For example, symptoms such as diplopia (Globas et al., 2008) or, more rarely, extrapyramidal signs such as bradykinesia or tremor (Bettencourt et al., 2011) may appear before gait disturbances. Three additional studies were available, but they focused on either the expanded protein alone (Hübener-Schmid et al., 2021; Prudencio et al., 2020), or the total ataxin-3 protein (Gonsior et al., 2021), limiting direct comparisons.

Extensive research has focused on the influence of intragenic variants and haplotypes of *ATXN3* on various aspects of MJD, including intergenerational repeat instability (Martins et al., 2008, 2014), somatic mosaicism (Sidky et al., 2024), age at disease onset (Melo et al., 2022), ataxia severity (SARA scores), non-ataxic manifestations (INAS score; (Elter et al., 2025) and ataxin-3 levels or physiological function (Harris et al., 2010; Johnson et al., 2019; Weishäupl et al., 2019). Our study, however, was the first to analyze the impact of genetic variants and haplotype on allele-specific transcript levels of *ATXN3*. Notably, the opposite trend of expression ratio observed in MJD expansion carriers from the Machado or the Joseph lineage suggested that *cis*-regulatory elements (inherited in a haplotype-dependent manner) influence allele-specific expression. This seems to be maintained at the protein level (Prudencio et al., 2020), although findings should be interpreted with caution given the very small sample size (fibroblast cells from one MJD patient carrying the rs7158733_C allele *versus* four patients carrying the rs7158733_A allele).

Overall, the imbalance *ATXN3* ASE does not seem to correlate with disease features such as the size of CAG tract or the age at onset. However, a more detailed examination of the distributions of these variables revealed several important findings. Although the expanded allele was more highly expressed in carriers of the Joseph lineage, which would typically predict a more severe phenotype (*i.e.,* earlier onset), these individuals tended to have lower non-expanded and expanded alleles, and therefore a later predicted age at onset compared with Machado carriers. Despite these differences, the reported ages at onset were similar between the two lineages, suggesting that higher expression of the expanded allele may shift a greater proportion of Joseph-lineage carriers toward earlier onset, whereas higher expression of the non-expanded allele may contribute to a greater proportion of Machado carriers reporting later onset.

The effect of SNV-based haplotypes that distinguish MJD lineages on disease phenotypes was recently investigated using a larger cohort of MJD expansion carriers (Elter et al., 2025). In that study, the authors analyzed haplotypes defined by the rs12895357 and rs7158733 variants in 319 DNA samples from the ESMI MJD cohort. Overall, our results are consistent with the findings reported by Elter and colleagues (Elter et al., 2025). MJD patients carrying the haplotype rs12895357_C + rs7158733_A alleles (Joseph lineage, n=189) and those with the rs12895357_G + rs7158733_C alleles (Machado lineage, n=66) had similar CAG expanded repeat lengths (∼70 repeats) and comparable ages at onset (∼41 years). Median SARA of 9 points (ataxia severity) and median inventory of non-ataxia signs (INAS) score of ∼ 4.5 points were also similar (Elter et al., 2025). Nevertheless, the frequency of specific clinical signs, namely extensor plantar reflex, spasticity, and dystonia was significantly higher in patients from the Machado lineage compared to those carrying the Joseph lineage (Elter et al., 2025). It is likely that the higher frequency of these signs, such as dystonia, is associated with a higher frequency of larger expanded alleles, although this interpretation is limited by the lack of available genetic and clinical variable distributions.

Among the four SNVs defining the haplotypes associated with MJD lineages, rs7158733 is likely to have the greatest functional impact, as the C-to-A substitution introduces a nonsense mutation that generates a premature stop codon, producing a shorter ataxin-3 isoform. In a previous study, ataxin-3 knockout cells were produced to express only one of the three transcripts *ATXN3-251*, *ATXN3-214* carrying the rs7158733 C allele or *ATXN3-214* carrying the A allele, each in *cis* with either the non-expanded or expanded CAG tract, thereby producing the corresponding proteins isoforms, named ataxin-3c, ataxin-3aL and ataxin-3aS, respectively (Weishäupl et al., 2019). In this context, MJD expansion carriers of the Joseph lineage produce the expanded ataxin-3aS isoform, which, compared with the expanded ataxin-3aL isoform (Machado lineage), exhibited similar steady-state levels, but faster degradation through both autophagy and proteasomal pathways, a higher propensity for nuclear rather than cytoplasmic localization, and distinct interaction networks. In terms of aggregation behavior, expanded ataxin-3aS formed fewer but larger aggregates, whereas expanded ataxin-3aL formed multiple smaller aggregates (Weishäupl et al., 2019). At the protein level, differences between the two ataxin-3 isoforms may contribute to MJD phenotypic variability; however, a direct comparison with such findings was not possible because our analysis quantified total *ATXN3* RNA rather than specific transcripts (e.g., *ATXN3-214*). In fact, the transcript most likely contributing to the majority of total *ATXN3* RNA measured in blood is the *ATXN3-251*, the canonical transcript (Raposo et al., 2024). SNVs rs7158733 and rs16999141 are also predicted to activate a cryptic acceptor splice site and alter the ESE/ESS motifs ratio (−10), respectively, in three out of four transcripts most abundantly expressed in blood, *ATXN3-251, ATXN3-214* and *ATXN3-254*. Both predictions suggest the potential for aberrant splicing, but additional functional studies are needed to experimentally confirm these effects.

In summary, we developed a novel protocol to assess the allele-specific expression of *ATXN3*. Our findings show that *ATXN3* exhibits allelic imbalance, notably haplotype-dependent, underlying the importance of determining MJD lineages. This imbalance, however, may also be tissue-specific, highlighting the need for future studies using brain samples. In fact, the coexistence of tissue-specific *cis*-regulation and allele-specific regulation has been previously observed (*e.g.*, Zhang et al., 2009). Deeper research of the RNA biology of expanded *ATXN3* transcripts should provide valuable insight into the pathogenesis of MJD.

## DECLARATIONS

### Availability of data and materials

Data is available in this manuscript and in supplementary material.

## Author contributions

Design and conceptualization of the study: MR, ML; Subject recruitment/Acquisition of participants data: MR, SP, AFF, ARVM, LT, JS, SM, ML; Acquisition of molecular data: MR, SP, SM, AMS; Statistical analysis of data: MR, SM; Drafting of the manuscript: ARVM, SM, MR, ML; Revision of the manuscript: MR, SP, AFF, ARVM, LT, JS, SM, ML, DGM, AMS. All authors read and approved the final manuscript.

## Data Availability

All data produced in the present work are contained in the manuscript.

## Acknowledgements

We thank all MJD subjects and their families for their participation.

## REFERENCES

Akçimen, F., Martins, S., Liao, C., Bourassa, C. V, Catoire, H., Nicholson, G. A., Riess, O., Raposo, M., França, M. C., Vasconcelos, J., Lima, M., Lopes-Cendes, I., Saraiva-Pereira, M. L., Jardim, L. B., Sequeiros, J., Dion, P. A., & Rouleau, G. A. (2020). Genome-wide association study identifies genetic factors that modify age at onset in Machado-Joseph disease. Aging, 12(6), 4742–4756. 10.18632/aging.102825

Bettencourt, C., & Lima, M. (2011). Machado-Joseph Disease: from first descriptions to new perspectives. Orphanet Journal of Rare Diseases, 6, 35. 10.1186/1750-1172-6-35

Bettencourt, C., Santos, C., Coutinho, P., Rizzu, P., Vasconcelos, J., Kay, T., Cymbron, T., Raposo, M., Heutink, P., & Lima, M. (2011). Parkinsonian phenotype in Machado-Joseph disease (MJD/SCA3): A two-case report. BMC Neurology, 11. 10.1186/1471-2377-11-131

Breuer, P., Rasche, T., Han, X., Faber, J., Haustein, K., Klockgether, T., & Wüllner, U. (2022). The Ratio of Expanded to Normal Ataxin 3 in Peripheral Blood Mononuclear Cells Correlates with the Age at Onset in Spinocerebellar Ataxia Type 3. In Movement disorders: official journal of the Movement Disorder Society (Vol. 37, Number 5, pp. 1098–1099). 10.1002/mds.28962

Castel, S. E., Aguet, F., Mohammadi, P., Ardlie, K. G., & Lappalainen, T. (2020). A vast resource of allelic expression data spanning human tissues. Genome Biology, 21(1), 234. 10.1186/s13059-020-02122-z

Costa, I. P. D., Almeida, B. C., Sequeiros, J., Amorim, A., & Martins, S. (2019). A pipeline to assess disease-associated haplotypes in repeat expansion disorders: The example of MJD/SCA3 locus. Frontiers in Genetics, 10(FEB), 1–9. 10.3389/fgene.2019.00038

Coutinho, P. (1992). Doença de Machado-Joseph: Tentativa de definição. Instituto de Ciências Biomédicas de Abel Salazar da Universidade do Porto.

Da Silva, J. D., Teixeira-Castro, A., & Maciel, P. (2019). From Pathogenesis to Novel Therapeutics for Spinocerebellar Ataxia Type 3: Evading Potholes on the Way to Translation. Neurotherapeutics, 16(4), 1009–1031. 10.1007/s13311-019-00798-1

de Mattos, E. P., Leotti, V. B., Soong, B.-W., Raposo, M., Lima, M., Vasconcelos, J., Fussiger, H., Souza, G. N., Kersting, N., Furtado, G. V, Saute, J. A. M., Camey, S. A., Saraiva-Pereira, M. L., & Jardim, L. B. (2019). Age at onset prediction in spinocerebellar ataxia type 3 changes according to population of origin. European Journal of Neurology, 26(1), 113–120. 10.1111/ene.13779

Desmet, F.-O., Hamroun, D., Lalande, M., Collod-Béroud, G., Claustres, M., & Béroud, C. (2009). Human Splicing Finder: an online bioinformatics tool to predict splicing signals. Nucleic Acids Research, 37(9), e67. 10.1093/nar/gkp215

Dyer, S. C., Austine-Orimoloye, O., Azov, A. G., Barba, M., Barnes, I., Barrera-Enriquez, V. P., Becker, A., Bennett, R., Beracochea, M., Berry, A., Bhai, J., Bhurji, S. K., Boddu, S., Branco Lins, P. R., Brooks, L., Ramaraju, S. B., Campbell, L. I., Martinez, M. C., Charkhchi, M., … Yates, A. D. (2025). Ensembl 2025. Nucleic Acids Research, 53(D1), D948–D957. 10.1093/nar/gkae1071

Elter, T. L., Sturm, D., Santana, M. M., Schaprian, T., Raposo, M., Melo, A. R. V., Lima, M., Koyak, B., Oender, D., Grobe-Einsler, M., Lopes, S., Silva, P., de Almeida, L. P., Giunti, P., Garcia-Moreno, H., Nethisinhe, S., de Vries, J., van de Warrenburg, B. P., van Gaalen, J., … Hübener-Schmid, J. (2025). Regional distribution of polymorphisms associated to the disease-causing gene of spinocerebellar ataxia type 3. Journal of Neurology, 272(1). 10.1007/s00415-024-12829-9

Evers, M. M., Toonen, L. J. A., & van Roon-Mom, W. M. C. (2013). Ataxin-3 Protein and RNA Toxicity in Spinocerebellar Ataxia Type 3: Current Insights and Emerging Therapeutic Strategies. Molecular Neurobiology. 10.1007/s12035-013-8596-2

Gaspar, C., Lopes-Cendes, I., DeStefano, A. L., Maciel, P., Silveira, I., Coutinho, P., MacLeod, P., Sequeiros, J., Farrer, L. A., & Rouleau, G. A. (1996). Linkage disequilibrium analysis in Machado-Joseph disease patients of different ethnic origins. Human Genetics, 98(5), 620–624. 10.1007/s004390050270

Globas, C., du Montcel, S. T., Baliko, L., Boesch, S., Depondt, C., DiDonato, S., Durr, A., Filla, A., Klockgether, T., Mariotti, C., Melegh, B., Rakowicz, M., Ribai, P., Rola, R., Schmitz-Hubsch, T., Szymanski, S., Timmann, D., Van de Warrenburg, B. P., Bauer, P., & Schols, L. (2008). Early symptoms in spinocerebellar ataxia type 1, 2, 3, and 6. Movement Disorders : Official Journal of the Movement Disorder Society, 23(15), 2232–2238. 10.1002/mds.22288

Gonsior, K., Kaucher, G. A., Pelz, P., Schumann, D., Gansel, M., Kuhs, S., Klockgether, T., Forlani, S., Durr, A., Hauser, S., Rattay, T. W., Synofzik, M., Hengel, H., Schöls, L., Rieß, O. H., & Hübener-Schmid, J. (2021). PolyQ-expanded ataxin-3 protein levels in peripheral blood mononuclear cells correlate with clinical parameters in SCA3: a pilot study. Journal of Neurology, 268(4), 1304–1315. 10.1007/s00415-020-10274-y

Goto, J., Watanabe, M., Ichikawa, Y., Yee, S. B., Ihara, N., Endo, K., Igarashi, S., Takiyama, Y., Gaspar, C., Maciel, P., Tsuji, S., Rouleau, G. A., & Kanazawa, I. (1997). Machado-Joseph disease gene products carrying different carboxyl termini. Neuroscience Research, 28(4), 373–377. 10.1016/s0168-0102(97)00056-4

Harris, G. M., Dodelzon, K., Gong, L., Gonzalez-Alegre, P., & Paulson, H. L. (2010). Splice isoforms of the polyglutamine disease protein ataxin-3 exhibit similar enzymatic yet different aggregation properties. PLoS ONE, 5(10). 10.1371/journal.pone.0013695

Hübener-Schmid, J., Kuhlbrodt, K., Peladan, J., Faber, J., Santana, M. M., Hengel, H., Jacobi, H., Reetz, K., Garcia-Moreno, H., Raposo, M., van Gaalen, J., Infante, J., Steiner, K. M., de Vries, J., Verbeek, M. M., Giunti, P., Pereira de Almeida, L., Lima, M., van de Warrenburg, B., … Riess, O. (2021). Polyglutamine-Expanded Ataxin-3: A Target Engagement Marker for Spinocerebellar Ataxia Type 3 in Peripheral Blood. Movement Disorders, 36(11), 2675–2681. 10.1002/mds.28749

Jacobi, H., Rakowicz, M., Rola, R., Fancellu, R., Mariotti, C., Charles, P., Dürr, A., Küper, M., Timmann, D., Linnemann, C., Schöls, L., Kaut, O., Schaub, C., Filla, A., Baliko, L., Melegh, B., Kang, J.-S., Giunti, P., van de Warrenburg, B. P. C., … Klockgether, T. (2013). Inventory of Non-Ataxia Signs (INAS): Validation of a New Clinical Assessment Instrument. The Cerebellum, 12(3), 418–428. 10.1007/s12311-012-0421-3

Joachimiak, P., Ciesiołka, A., Kozłowska, E., Świtoński, Paweł M, Figura, G., Ciołak, A., Adamek, G., Surdyka, M., Kalinowska-Pośka, Ż., Figiel, M., Caron, N. S., Hayden, M. R., & Fiszer, A. (2023). Allele-specific quantitation of ATXN3 and HTT transcripts in polyQ disease models. BMC Biology, 21(1), 17. 10.1186/s12915-023-01515-3

Johnson, S. L., Blount, J. R., Libohova, K., Ranxhi, B., Paulson, H. L., Tsou, W.-L., & Todi, S. V. (2019). Differential toxicity of ataxin-3 isoforms in Drosophila models of Spinocerebellar Ataxia Type 3. Neurobiology of Disease, 132, 104535. 10.1016/j.nbd.2019.104535

Lima, M., Raposo, M., Ferreira, A., Melo, A. R. V., Pavão, S., Medeiros, F., Teves, L., Gonzalez, C., Lemos, J., Pires, P., Lopes, P., Valverde, D., Gonzalez, J., Kay, T., & Vasconcelos, J. (2023). The Homogeneous Azorean Machado-Joseph Disease Cohort: Characterization and Contributions to Advances in Research. Biomedicines, 11(2). 10.3390/biomedicines11020247

Liu, W., Chaurette, J., Pfister, E. L., Kennington, L. A., Chase, K. O., Bullock, J., Vonsattel, J. P. G., Faull, R. L. M., Macdonald, D., DiFiglia, M., Zamore, P. D., & Aronin, N. (2013). Increased Steady-State Mutant Huntingtin mRNA in Huntington’s Disease Brain. Journal of Huntington’s Disease, 2(4), 491–500. 10.3233/JHD-130079

Maciel, P., Gaspar, C., Guimarães, L., Goto, J., Lopes-Cendes, I., Hayes, S., Arvidsson, K., Dias, A., Sequeiros, J., Sousa, A., & Rouleau, G. A. (1999). Study of three intragenic polymorphisms in the Machado-Joseph disease gene (MJD1) in relation to genetic instability of the (CAG)n tract. European Journal of Human Genetics : EJHG, 7(2), 147–156. 10.1038/sj.ejhg.5200264

Martins, S., Calafell, F., Gaspar, C., Wong, V. C. N., Silveira, I., Nicholson, G. A., Brunt, E. R., Tranebjaerg, L., Stevanin, G., Hsieh, M., Soong, B., Loureiro, L., Dürr, A., Tsuji, S., Watanabe, M., Jardim, L. B., Giunti, P., Riess, O., Ranum, L. P. W., … Sequeiros, J. (2007). Asian Origin for the Worldwide-Spread Mutational Event in Machado-Joseph Disease. Archives of Neurology, 64(10), 1502. 10.1001/archneur.64.10.1502

Martins, S., Coutinho, P., Silveira, I., Giunti, P., Jardim, L. B., Calafell, F., Sequeiros, J., & Amorim, A. (2008). Cis -acting factors promoting the CAG intergenerational instability in Machado–Joseph disease. American Journal of Medical Genetics Part B: Neuropsychiatric Genetics, 147B(4), 439–446. 10.1002/ajmg.b.30624

Martins, S., Pearson, C. E., Coutinho, P., Provost, S., Amorim, A., Dubé, M.-P., Sequeiros, J., & Rouleau, G. A. (2014). Modifiers of (CAG)n instability in Machado–Joseph disease (MJD/SCA3) transmissions: an association study with DNA replication, repair and recombination genes. Human Genetics, 133(10), 1311–1318. 10.1007/s00439-014-1467-8

Martins, S., & Sequeiros, J. (2018). Origins and Spread of Machado-Joseph Disease Ancestral Mutations Events (pp. 243–254). 10.1007/978-3-319-71779-1_12

Martins, S., Soong, B.-W., Wong, V. C. N., Giunti, P., Stevanin, G., Ranum, L. P. W., Sasaki, H., Riess, O., Tsuji, S., Coutinho, P., Amorim, A., Sequeiros, J., & Nicholson, G. A. (2012). Mutational origin of Machado-Joseph disease in the Australian Aboriginal communities of Groote Eylandt and Yirrkala. Archives of Neurology, 69(6), 746–751. 10.1001/archneurol.2011.2504

McLaren, W., Gil, L., Hunt, S. E., Riat, H. S., Ritchie, G. R. S., Thormann, A., Flicek, P., & Cunningham, F. (2016). The Ensembl Variant Effect Predictor. Genome Biology, 17(1), 122. 10.1186/s13059-016-0974-4

Melo, A. R. V., Raposo, M., Ventura, M., Martins, S., Pavão, S., Alonso, I., Bettencourt, C., & Lima, M. (2022). Genetic Variation in ATXN3 (Ataxin-3) 3′UTR: Insights into the Downstream Regulatory Elements of the Causative Gene of Machado-Joseph Disease/Spinocerebellar Ataxia Type 3. The Cerebellum. 10.1007/s12311-021-01358-0

Nalavade, R., Griesche, N., Ryan, D. P., Hildebrand, S., & Krauß, S. (2013). Mechanisms of RNA-induced toxicity in CAG repeat disorders. Cell Death & Disease, 4(8), e752–e752. 10.1038/cddis.2013.276

Nishiyama, K., Murayama, S., Goto, J., Watanabe, M., Hashida, H., Kanazawa, I., Katayama, S., Nakamura, S., & Nomura, Y. (1996). Regional and cellular expression of the machado-joseph disease gene in brains of normal and affected individuals. Annals of Neurology, 40(5), 776–781. 10.1002/ana.410400514

Ogun, S. A., Martins, S., Adebayo, P. B., Dawodu, C. O., Sequeiros, J., & Finkel, M. F. (2015). Machado-Joseph disease in a Nigerian family: mutational origin and review of the literature. European Journal of Human Genetics: EJHG, 23(2), 271–273. 10.1038/ejhg.2014.77

Prudencio, M., Garcia-Moreno, H., Jansen-West, K. R., AL-Shaikh, R. H., Gendron, T. F., Heckman, M. G., Spiegel, M. R., Carlomagno, Y., Daughrity, L. M., Song, Y., Dunmore, J. A., Byron, N., Oskarsson, B., Nicholson, K. A., Staff, N. P., Gorcenco, S., Puschmann, A., Lemos, J., Januário, C., … Petrucelli, L. (2020). Toward allele-specific targeting therapy and pharmacodynamic marker for spinocerebellar ataxia type 3. Science Translational Medicine, 12(566). 10.1126/scitranslmed.abb7086

Raposo, M., Bettencourt, C., Maciel, P., Gao, F., Ramos, A., Kazachkova, N., Vasconcelos, J., Kay, T., Rodrigues, A. J., Bettencourt, B., Bruges-Armas, J., Geschwind, D., Coppola, G., & Lima, M. (2015). Novel candidate blood-based transcriptional biomarkers of Machado-Joseph disease. Movement Disorders, 30(7), 968–975. 10.1002/mds.26238

Raposo, M., Bettencourt, C., Melo, A. R. V., Ferreira, A. F., Alonso, I., Silva, P., Vasconcelos, J., Kay, T., Saraiva-Pereira, M. L., Costa, M. D., Vilasboas-Campos, D., Bettencourt, B. F., Bruges-Armas, J., Houlden, H., Heutink, P., Jardim, L. B., Sequeiros, J., Maciel, P., & Lima, M. (2021). Novel Machado-Joseph disease-modifying genes and pathways identified by whole-exome sequencing. Neurobiology of Disease, 162, 105578. 10.1016/j.nbd.2021.105578

Raposo, M., Hübener-Schmid, J., Tagett, R., Ferreira, Ana F, Vieira Melo, A. R., Vasconcelos, J., Pires, P., Kay, T., Garcia-Moreno, H., Giunti, P., Santana, M. M., Pereira de Almeida, L., Infante, J., van de Warrenburg, B. P., de Vries, J. J., Faber, J., Klockgether, T., Casadei, N., Admard, J., … Lima, M. (2024). Blood and cerebellar abundance of ATXN3 splice variants in spinocerebellar ataxia type 3/Machado-Joseph disease. Neurobiology of Disease, 193, 106456. 10.1016/j.nbd.2024.106456

Raposo, M., Ramos, A., Bettencourt, C., & Lima, M. (2015). Replicating studies of genetic modifiers in spinocerebellar ataxia type 3: can homogeneous cohorts aid? Brain, 138(12), e398–e398. 10.1093/brain/awv206

Rohilla, K. J., & Gagnon, K. T. (2017). RNA biology of disease-associated microsatellite repeat expansions. Acta Neuropathologica Communications, 5(1), 63. 10.1186/s40478-017-0468-y

Santos, C., Malheiro, S., Correia, M., & Damásio, J. (2023). Gene Suppression Therapies in Hereditary Cerebellar Ataxias: A Systematic Review of Animal Studies. Cells, 12(7), 1037. 10.3390/cells12071037

Schmitz-Hübsch, T., du Montcel, S. T., Baliko, L., Berciano, J., Boesch, S., Depondt, C., Giunti, P., Globas, C., Infante, J., Kang, J.-S., Kremer, B., Mariotti, C., Melegh, B., Pandolfo, M., Rakowicz, M., Ribai, P., Rola, R., Schöls, L., Szymanski, S., … Klockgether, T. (2006). Scale for the assessment and rating of ataxia. Neurology, 66(11), 1717–1720. 10.1212/01.wnl.0000219042.60538.92

Sharony, R., Martins, S., Costa, I. P. D., Zaltzman, R., Amorim, A., Sequeiros, J., & Gordon, C. R. (2019). Yemenite-Jewish families with Machado-Joseph disease (MJD/SCA3) share a recent common ancestor. European Journal of Human Genetics: EJHG, 27(11), 1731–1737. 10.1038/s41431-019-0449-7

Sidky, Ahmed M., Melo, A. R. V., Kay, T. T., Raposo, M., Lima, M., & Monckton, D. G. (2024). Age-dependent somatic expansion of the ATXN3 CAG repeat in the blood and buccal swab DNA of individuals with spinocerebellar ataxia type 3/Machado-Joseph disease. Human Genetics, 143(11), 1363–1378. 10.1007/s00439-024-02698-7

St. Pierre, C. L., Macias-Velasco, J. F., Wayhart, J. P., Yin, L., Semenkovich, C. F., & Lawson, H. A. (2022). Genetic, epigenetic, and environmental mechanisms govern allele-specific gene expression. Genome Research, 32(6), 1042–1057. 10.1101/gr.276193.121

Vogel, C., & Marcotte, E. M. (2012). Insights into the regulation of protein abundance from proteomic and transcriptomic analyses. Nature Reviews. Genetics, 13(4), 227–232. 10.1038/nrg3185

Weber, J. J., Sowa, A. S., Binder, T., & Hübener, J. (2014). From Pathways to Targets: Understanding the Mechanisms behind Polyglutamine Disease. BioMed Research International, 2014, 1–22. 10.1155/2014/701758

Weishäupl, D., Schneider, J., Peixoto Pinheiro, B., Ruess, C., Dold, S. M., von Zweydorf, F., Gloeckner, C. J., Schmidt, J., Riess, O., & Schmidt, T. (2019). Physiological and pathophysiological characteristics of ataxin-3 isoforms. Journal of Biological Chemistry, 294(2), 644–661. 10.1074/jbc.RA118.005801

Wu, Y., Peng, Y., & Wang, Y. (2015). An insight into advances in the pathogenesis and therapeutic strategies of spinocerebellar ataxia type 3. Reviews in the Neurosciences, 26(1). 10.1515/revneuro-2014-0040

Zhang, K., Li, J. B., Gao, Y., Egli, D., Xie, B., Deng, J., Li, Z., Lee, J.-H., Aach, J., Leproust, E. M., Eggan, K., & Church, G. M. (2009). Digital RNA allelotyping reveals tissue-specific and allele-specific gene expression in human. Nature Methods, 6(8), 613–618. 10.1038/nmeth.1357

