## Supplemental Material for "Allele-specific expression of *ATXN3* in blood samples of Machado-Joseph disease expansion carriers"

Melo *et al*.

*Contains:*

- Description of the protocol to determine allele-specific expression of the *ATXN3* (13 pages)
- Supplementary Figures: 1

**Comprehensive protocol for assessing allele-specific expression (ASE) of *ATXN3***

---

The protocol consists of two main steps: (1) identification of subjects heterozygous for rs1048755 and quantification of allele-specific transcripts using qPCR, and (2) determination of SNV–$(CAG)_n$ phasing through electrophoresis and Sanger sequencing of DNA fragments corresponding to the expanded and non-expanded *ATXN3* alleles. Prior to these core steps, three preparatory procedures were conducted to evaluate key methodological variables.

I. Preparatory procedures

(A) SNVs distribution in *ATXN3* transcripts

*ATXN3* produces at least 54 transcripts by alternative splicing (1,2). To guarantee the highest levels of allele-specific expression, the presence and location in the gene (coding exons, intron, UTRs) of the SNVs in the 54 transcripts were investigated (3). The distribution of SNVs across the 54 *ATXN3* transcripts is shown in Table 1. Several SNVs are in the intronic regions of the gene, which may limit the selection of SNVs suitable for analysis of allele-specific expression. Of the 12 analyzed SNVs, none are present in nine transcripts (Table 1).

Additionally, the abundance of *ATXN3* transcripts in blood was considered (2). Only eight transcripts are commonly expressed in blood (2). Moreover, none are present in four of the eight most abundant blood transcripts. However, four blood-abundant transcripts contain rs16999141, rs1048755 and rs12895357. The remaining SNVs are present in only one or two transcripts (Table 1). Given this, rs16999141, rs1048755, and rs12895357 have been selected for further use.

**Table 1.** Distribution of SNVs across the 54 *ATXN3* transcripts. Data were obtained from Ensembl (1). Transcripts highlighted in red are frequently expressed in blood samples (2). P=position of the SNV in the transcript. Consequence=consequence type of the SNV in the transcript.

| | | | rs16999141 | | rs1048755 | | rs12895357 | | rs7158733 | | rs11628764 | | rs10151135 | | rs3092822 | | rs709930 | | rs1055996 | | rs55966267 | | rs910369 | | rs7158238 | |
|---|---|---|---|---|---|---|---|---|---|---|---|---|---|---|---|---|---|---|---|---|---|---|---|---|---|---|
| Transcript ID | Name | bp | Consequence | P | Consequence | P | Consequence | P | Consequence | P | Consequence | P | Consequence | P | Consequence | P | Consequence | P | Consequence | P | Consequence | P | Consequence | P | Consequence | P |
| **Protein coding** | | | | | | | | | | | | | | | | | | | | | | | | | | |
| ENST00000644486.2 | *ATXN3-251* | 6884 | synonymous | 522 | missense | 664 | missense | 946 | intron | | 3 prime UTR | 380 | 3 prime UTR | 1229 | intron | | 3 prime UTR | 1267 | 3 prime UTR | 4381 | 3 prime UTR | 2222 | 3 prime UTR | 1498 | intron | |
| ENST00000340660.10 | *ATXN3-201* | 1205 | synonymous | 385 | missense | 527 | missense | 809 | intron | | | | 3 prime UTR | 1092 | intron | | 3 prime UTR | 1130 | | | | | | | intron | |
| ENST00000393287.9 | *ATXN3-203* | 6770 | synonymous | 408 | missense | 550 | missense | 832 | intron | | 3 prime UTR | 368 | 3 prime UTR | 1115 | intron | | 3 prime UTR | 1153 | 3 prime UTR | 4267 | 3 prime UTR | 2108 | 3 prime UTR | 1384 | intron | |
| ENST00000429774.6 | *ATXN3-204* | 6713 | synonymous | 351 | missense | 493 | missense | 775 | intron | | 3 prime UTR | 363 | 3 prime UTR | 1058 | intron | | 3 prime UTR | 1096 | 3 prime UTR | 4210 | 3 prime UTR | 2051 | 3 prime UTR | 1327 | intron | |
| ENST00000502250.5 | *ATXN3-206* | 1259 | 5 prime UTR | 226 | missense | 368 | missense | 650 | intron | | | | 3 prime UTR | 933 | intron | | 3 prime UTR | 971 | | | | | 3 prime UTR | 1202 | intron | |
| ENST00000503767.5 | *ATXN3-207* | 1315 | synonymous | 495 | missense | 637 | missense | 919 | intron | | | | 3 prime UTR | 1202 | intron | | 3 prime UTR | 1240 | | | | | | | intron | |
| ENST00000506466.5 | *ATXN3-209* | 637 | synonymous | 315 | missense | 457 | | | | | | | | | | | | | | | | | | | | |
| ENST00000526872.3 | *ATXN3-213* | 604 | splice region | | missense | 506 | | | | | | | | | | | | | | | | | | | | |
| ENST00000532032.5 | *ATXN3-214* | 1191 | synonymous | 502 | missense | 644 | missense | 926 | stop gained | 1057 | | | | | 3 prime UTR | 1117 | | | | | | | | | 3 prime UTR | 1175 |
| ENST00000545170.5 | *ATXN3-215* | 6950 | synonymous | 561 | missense | 703 | missense | 101 | intron | | 3 prime UTR | 386 | 3 prime UTR | 1295 | intron | | 3 prime UTR | 1333 | 3 prime UTR | 4447 | 3 prime UTR | 2288 | 3 prime UTR | 1564 | intron | |
| ENST00000553491.5 | *ATXN3-219* | 857 | synonymous | 361 | missense | 503 | missense | 785 | | | | | | | | | | | | | | | | | | |
| ENST00000554592.5 | *ATXN3-227* | 1010 | synonymous | 511 | missense | 653 | missense | 935 | | | | | | | | | | | | | | | | | | |
| ENST00000554672.6 | *ATXN3-228* | 683 | 5 prime UTR | 260 | missense | 402 | | | | | | | | | | | | | | | | | | | | |
| ENST00000555381.5 | *ATXN3-231* | 803 | synonymous | 304 | missense | 446 | missense | 728 | | | | | | | | | | | | | | | | | | |
| ENST00000556220.5 | *ATXN3-235* | 692 | synonymous | 196 | missense | 338 | missense | 620 | | | | | | | | | | | | | | | | | | |
| ENST00000557311.6 | *ATXN3-246* | 559 | 5 prime UTR | 63 | missense | 205 | missense | 487 | | | | | | | | | | | | | | | | | | |
| **Nonsense mediated decay** | | | | | | | | | | | | | | | | | | | | | | | | | | |
| ENST00000359366.10 | *ATXN3-202* | 2572 | 3 prime UTR | 444 | 3 prime UTR | 586 | 3 prime UTR | 868 | intron | | | | 3 prime UTR | 1151 | intron | | 3 prime UTR | 1189 | | | 3 prime UTR | 2144 | 3 prime UTR | 1420 | intron | |
| ENST00000515746.6 | *ATXN3-212* | 618 | missense | 261 | 3 prime UTR | 403 | | | | | | | | | | | | | | | | | | | | |
| ENST00000553488.5 | *ATXN3-218* | 1065 | 3 prime UTR | 515 | 3 prime UTR | 657 | 3 prime UTR | 993 | | | | | | | | | | | | | | | | | | |
| ENST00000553570.5 | *ATXN3-221* | 757 | missense | 261 | 3 prime UTR | 403 | 3 prime UTR | 685 | | | | | | | | | | | | | | | | | | |
| ENST00000554350.5 | *ATXN3-225* | 943 | 3 prime UTR | 447 | 3 prime UTR | 589 | 3 prime UTR | 871 | | | | | | | | | | | | | | | | | | |
| ENST00000554673.5 | *ATXN3-229* | 981 | 3 prime UTR | 601 | 3 prime UTR | 743 | | | | | | | | | | | | | | | | | | | | |
| ENST00000554994.5 | *ATXN3-230* | 922 | missense | 426 | 3 prime UTR | 568 | 3 prime UTR | 850 | | | | | | | | | | | | | | | | | | |
| ENST00000555816.5 | *ATXN3-232* | 760 | 3 prime UTR | 428 | intron | | 3 prime UTR | 685 | | | | | | | | | | | | | | | | | | |
| ENST00000556082.6 | *ATXN3-234* | 587 | 3 prime UTR | 428 | intron | | | | | | | | | | | | | | | | | | | | | |
| ENST00000556274.5 | *ATXN3-236* | 913 | splice region | | 3 prime UTR | 547 | 3 prime UTR | 841 | | | | | | | | | | | | | | | | | | |
| ENST00000556288.5 | *ATXN3-237* | 901 | splice region | | 3 prime UTR | 547 | 3 prime UTR | 829 | | | | | | | | | | | | | | | | | | |
| ENST00000556315.6 | *ATXN3-238* | 848 | 3 prime UTR | 425 | 3 prime UTR | 567 | | | | | | | | | | | | | | | | | | | | |
| ENST00000556374.5 | *ATXN3-240* | 1083 | 3 prime UTR | 533 | 3 prime UTR | 675 | 3 prime UTR | 101 | | | | | | | | | | | | | | | | | | |
| ENST00000556671.5 | *ATXN3-242* | 921 | splice region | | 3 prime UTR | 567 | 3 prime UTR | 849 | | | | | | | | | | | | | | | | | | |
| ENST00000556898.5 | *ATXN3-243* | 990 | splice region | | 3 prime UTR | 633 | 3 prime UTR | 915 | | | | | | | | | | | | | | | | | | |
| ENST00000556958.5 | *ATXN3-244* | 769 | missense | 273 | 3 prime UTR | 415 | 3 prime UTR | 697 | | | | | | | | | | | | | | | | | | |
| ENST00000557030.6 | *ATXN3-245* | 806 | synonymous | 314 | missense | 456 | 3 prime UTR | 734 | | | | | | | | | | | | | | | | | | |
| ENST00000646485.1 | *ATXN3-253* | 682 | 5 prime UTR | 263 | missense | 405 | | | | | | | | | | | | | | | | | | | | |
| ENST00000647161.1 | *ATXN3-254* | 1569 | synonymous | 520 | missense | 662 | missense | 944 | intron | | intron | - | intron | | intron | | intron | | intron | | intron | | intron | | intron | |
| **Protein coding CDS not defined** | | | | | | | | | | | | | | | | | | | | | | | | | | |
| ENST00000504047.1 | *ATXN3-208* | 426 | | | | | | | | | | | | | | | | | | | | | | | | |
| ENST00000511362.5 | *ATXN3-211* | 591 | | | | | | | | | | | | | | | | | | | | | | | | |
| ENST00000553287.5 | *ATXN3-216* | 726 | exon | 218 | exon | 360 | exon | 654 | | | | | | | | | | | | | | | | | | |
| ENST00000553309.5 | *ATXN3-217* | 1022 | exon | 514 | exon | 656 | exon | 950 | | | | | | | | | | | | | | | | | | |
| ENST00000553498.5 | *ATXN3-220* | 911 | exon | 361 | exon | 503 | exon | 839 | | | | | | | | | | | | | | | | | | |
| ENST00000553686.5 | *ATXN3-222* | 869 | exon | 361 | exon | 503 | exon | 797 | | | | | | | | | | | | | | | | | | |
| ENST00000554040.5 | *ATXN3-223* | 592 | exon | 263 | intron | | exon | 520 | | | | | | | | | | | | | | | | | | |
| ENST00000554214.5 | *ATXN3-224* | 771 | exon | 263 | exon | 405 | exon | 699 | | | | | | | | | | | | | | | | | | |
| ENST00000554491.5 | *ATXN3-226* | 1064 | exon | 514 | exon | 656 | exon | 992 | | | | | | | | | | | | | | | | | | |
| ENST00000555958.5 | *ATXN3-233* | 857 | exon | 349 | exon | 491 | exon | 785 | | | | | | | | | | | | | | | | | | |
| ENST00000556339.5 | *ATXN3-239* | 525 | intron | | intron | | exon | 453 | | | | | | | | | | | | | | | | | | |
| ENST00000556644.5 | *ATXN3-241* | 372 | intron | | intron | | exon | 300 | | | | | | | | | | | | | | | | | | |
| ENST00000564606.6 | *ATXN3-247* | 460 | | | | | | | | | | | | | | | | | | | | | | | | |
| ENST00000642417.1 | *ATXN3-249* | 1098 | | | | | | | | | | | | | | | | | | | | | | | | |
| ENST00000642896.1 | *ATXN3-250* | 423 | | | | | | | | | | | | | | | | | | | | | | | | |
| ENST00000644720.1 | *ATXN3-252* | 388 | | | | | | | | | | | | | | | | | | | | | | | | |
| **Retained intron** | | | | | | | | | | | | | | | | | | | | | | | | | | |
| ENST00000454964.3 | ATXN3-205 | 598 | | | | | | | | | | | | | | | | | | | | | | | | |
| ENST00000507965.5 | ATXN3-210 | 708 | | | | | | | | | | | | | | | | | | | | | | | | |
| ENST00000568290.1 | ATXN3-248 | 671 | | | | | | | | | | | | | | | | | | | | | | | | |

(B) Performance of predesigned TaqMan genotyping assays

Predesigned TaqMan genotyping assays for the most frequently observed SNVs, in the 54 *ATXN3* transcripts in blood (according to the findings from Preparatory Step A), were ordered for rs16999141, rs1048755, and rs12895357 (with the following IDs - C_16189421_10, C_34507655_10, and C_32097352_10, respectively), and the ability to accurately define the three genotype clusters (homozygote A, homozygote B, and heterozygote) was investigated. Quantitative real-time PCR (qPCR) for allelic discrimination was performed using 20 ng of DNA per reaction, along with the respective predesigned TaqMan genotyping assay and the TaqPath™ ProAmp™ Master Mix, according to the manufacturer's instructions. TaqMan genotyping reactions were run in an ABI StepOnePlus™ Real-Time PCR System (Applied Biosystems). Raw data were further analyzed in TaqMan Genotyper Software (Applied Biosystems), including generation of the allelic discrimination plots.

We successfully obtained the three genotype clusters for rs1048755, whereas the results for rs16999141 and rs12895357 were unsatisfactory (Figure 1a). Specifically, we were unable to successfully distinguish the three genotype clusters for rs12895357, likely due to limited amplification. The assay, which includes two primers and probes, was developed based on the reference genome sequence, which contains only 14 CAG repeats; however, in samples of MJD expansion carriers, typically having over 60 repeats, this expanded region interferes with primer/probe binding, resulting in a lack of amplification. Additionally, the assay for rs16999141 showed weaker fluorescence signals compared to rs1048755, although the conditions were the same for both assays, including the cDNA samples. Furthermore, homozygotes for the rs16999141 C allele were not observed as the T allele is in complete linkage disequilibrium (LD) with the CAG expansion, which can influence the accurate assignment of genotype clusters.

(C) Optimization of cDNA input

To adapt the use of cDNA, instead of genomic DNA (as TaqMan Genotyping assays are optimized for gDNA input), we improved the cDNA concentration, by testing four different amounts per reaction: 20 ng (the maximum concentration recommended by the TaqMan SNP Genotyping protocol for DNA input), 40 ng, 100 ng (the maximum of cDNA amount recommended by the TaqMan Gene Expression protocol) and 1000 ng (Table 2). qPCR experiments were conducted as previously described (Preparatory Step B).

**Table 2.** Cycle threshold (Ct) values from qPCR were obtained during preparatory steps to optimize cDNA input, testing four different input amounts per reaction. Twelve cDNA samples from carriers of MJD mutation were used. Und = undetermined.

| | cDNA | | | |
|---|---|---|---|---|
| | **20ng** | **40ng** | **100ng** | **1000ng** |
| S1 | 35.282<br>34.566 | 32.925<br>32.935 | 32.866<br>32.722 | 27.568<br>27.315 |
| S2 | 35.227<br>34.435 | 37.774<br>34.725 | 33.587<br>33.476 | 29.482<br>28.886 |
| S3 | | 35.574<br>34.619 | | 28.714<br>28.390 |
| S4 | und<br>37.148 | und<br>36.726 | 35.943<br>33.526 | |
| S5 | 36.988<br>35.822 | 35.330<br>35.284 | 35.118<br>34.348 | 28.567<br>28.169 |
| S6 | 37.787<br>und | | 33.396<br>32.314 | 29.050<br>28.697 |
| S7 | 38.071<br>und | | 32.529<br>32.081 | 28.152<br>27.810 |
| S8 | | 36.911<br>35.582 | 36.543<br>36.529 | 29.675<br>29.364 |
| S9 | | 36.702<br>38.361 | 32.622<br>36.017 | 28.021<br>31.028 |
| S10 | | | | 28.702<br>31.301 |
| S11 | | 35.193<br>35.003 | 34.195<br>33.939 | 29.230<br>28.797 |
| S12 | und<br>36.492 | | 35.862<br>31.785 | 32.278<br>28.574 |

The cycle thresholds obtained with 1000 ng of cDNA were the closest match to those achieved with the standard input of 20 ng of DNA (Figure 1b). The consistency of the cycle threshold values in intra and inter-runs was also assessed (data not shown); the variation across replicates was not significant and was similar either using DNA or cDNA.

II. Protocol development

Step 1: Identification of heterozygotes and quantification of allele-specific transcripts

Ninety cDNA blood samples of MJD subjects from the Azorean cohort were used. A predesigned TaqMan assay (ID - C_16189421_10) was ordered to identify heterozygotes for rs1048755. Two probes labelled either with VIC reporter dye or 6-FAM were selected against each of the mRNA variants (G or A), which were complementary to one mRNA but mismatched to the other. qPCR experiments were conducted as previously described in Preparatory Step B, but using the optimized cDNA concentration previously determined in Preparatory Step C. Cycle thresholds (Ct) of the mRNA with G variant and the mRNA with the A variant were retrieved from the SDS v.2.3 software (Applied Biosystems).

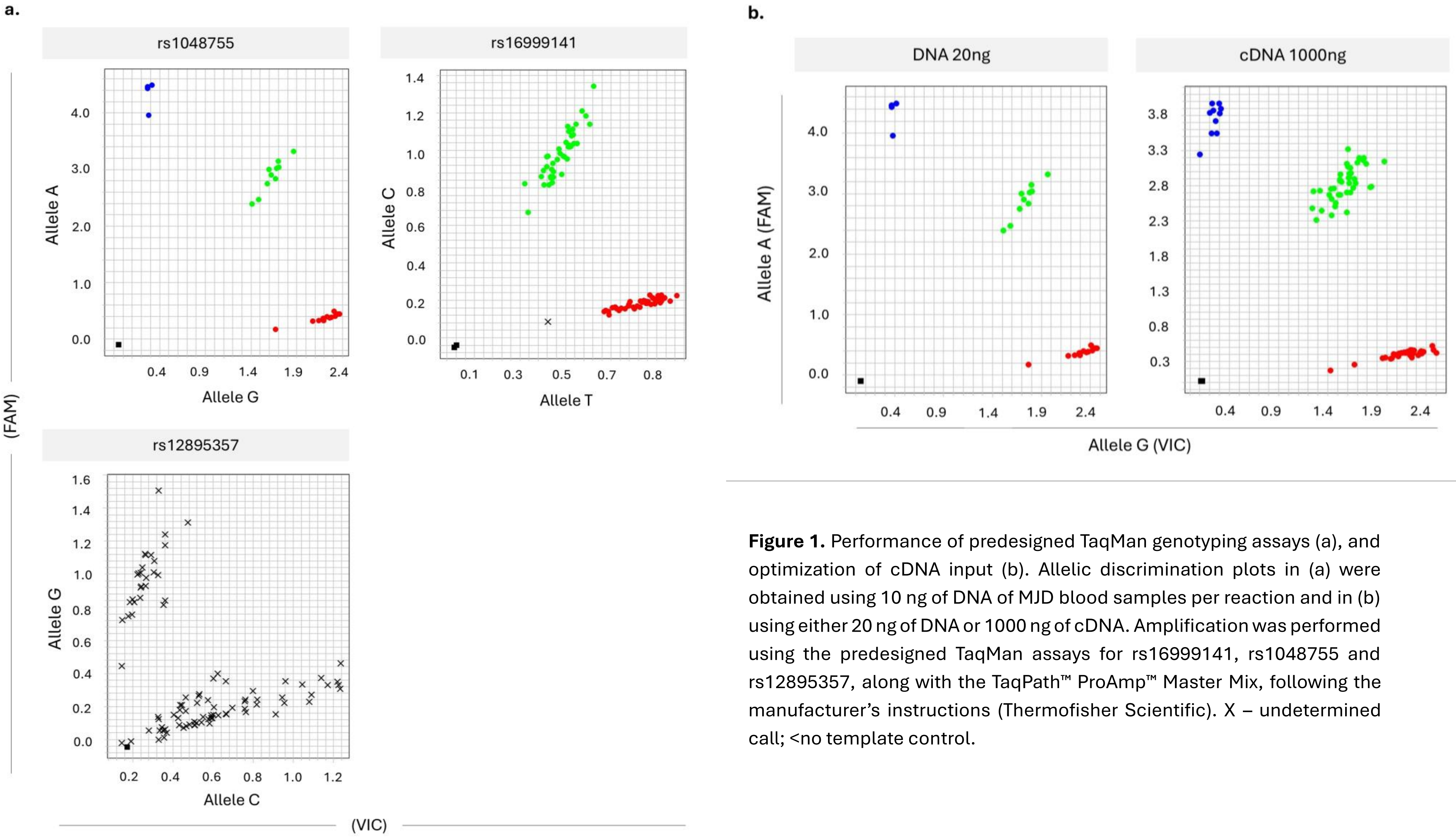


**Figure 1.** Performance of predesigned TaqMan genotyping assays (a), and optimization of cDNA input (b). Allelic discrimination plots in (a) were obtained using 10 ng of DNA of MJD blood samples per reaction and in (b) using either 20 ng of DNA or 1000 ng of cDNA. Amplification was performed using the predesigned TaqMan assays for rs16999141, rs1048755 and rs12895357, along with the TaqPath™ ProAmp™ Master Mix, following the manufacturer's instructions (Thermofisher Scientific). X – undetermined call; <no template control.

Step 1.1: Genetic variation of *ATXN3*

As the analysis of rs1048755 alone did not allow us to distinguish non-expanded and expanded alleles in all samples (*i.e.*, in samples from homozygotes for this SNV), the genotypes of other SNVs within the *ATXN3* were also analyzed. In samples from the Azorean cohort, genotypes for rs1048755 and rs16999141 (ENST00000644486.2: c.492C>T) were obtained by qPCR experiments, as previously described, but using a total of 10 ng of DNA per reaction. Genotypes for rs12895357 (ENST00000644486.2:c.916G>C),rs11628764(ENST00000644486.2:c.*2687A>G),rs10151135(ENST00000644486.2:c.*113C>A),rs3092822(ENST00000644486.2:c.991+116A>C),rs709930(ENST00000644486.2:c.*151G>A),rs1055996(ENST00000644486.2:c.*3265T>C),rs55966267(ENST00000644486.2:c.*1106A>G),rs910369(ENST00000644486.2:c.*382G>T), rs7158238 (ENST00000644486.2:c.991+174G>A) and rs7158733 (ENST00000644486.2:c.991+56C>A) were available from previous works (4–7). Genotype frequencies (Table 3) and linkage disequilibrium tests (Table 4) for the 12 SNVs were determined using Arlequin software v.3.5 (8). Importantly, two factors may influence heterozygosity levels of SNVs in MJD cohorts: (i) the frequency of alleles in *cis* with the CAG expansion (which may define the mutational lineage (Machado and Joseph), and (ii) the allele frequency of SNVs in the respective control population. The Machado lineage is more frequent in Mainland Portuguese MJD patients (~70%) whereas in the Azorean cohort both lineages have similar genotypic frequencies (Table 3).

Along with data from Portugal (mainland and Azores), SNV genotype and allele frequencies from control European and East Asian populations, which are the two major populations where MJD is most frequently reported, were retrieved from the 1000 Genomes Project (https://www.internationalgenome.org). A population differentiation exact test to compare genotypic frequencies from the Portuguese Azorean and mainland MJD cohorts, and from European and East Asian healthy populations was conducted. For the four SNVs rs16999141, rs1048755, rs12895357, rs7158733 for which genotype data from mainland Portugal and Azores was available (Table 3), similar genotype frequencies across the two cohorts were observed (differentiation exact test, p-value>0.05). For rs16999141, rs1048755, rs12895357 and rs7158733 differences were found between the mainland Portugal and Azores patients and European healthy population when compared to values of East Asian healthy population (differentiation exact test, p-value<0.05).

For the other SNVs a population differentiation exact test to compare genotypic frequencies from the Portuguese Azorean and European and East Asian healthy populations was also performed. In the cases of rs11628764, rs10151135, rs3092822 and rs55966267 different genotype frequencies across the three cohorts were observed (differentiation exact test, p-value<0.05). For rs709930, rs1055996 and rs910369 were also observed a difference in genotype frequencies across the Portuguese Azorean and European healthy population and between European healthy population and East Asian healthy population (differentiation exact test, p-value<0.05). For the same SNVs were observed similar frequencies across Portuguese Azorean population and East Asian healthy population (differentiation exact test, p-value>0.05). Finally, for SNV rs7158238 no analysis was performed due to the lack of data.

**Table 3.** Genotypes of *ATXN3* single nucleotide variants (SNVs) observed in (a) Azorean and (b) mainland Portuguese carriers of MJD mutation, grouped by Machado and Joseph lineages and (c) European and East Asian control populations. The allelic variants of all the 12 SNVs are shown in the anti-sense strand. Azorean MJD subjects from three families seem to belong to Machado or Joseph sub-lineages and were not considered in the frequencies.

**a.**

| | Portuguese Azorean MJD cohort | | | | | | | | | | | |
|---|---|---|---|---|---|---|---|---|---|---|---|---|
| | rs16999141 | rs1048755 | rs12895357 | rs7158733 | rs11628764 | rs10151135 | rs3092822 | rs709930 | rs1055996 | rs55966267 | rs910369 | rs7158238 |
| MJD patients, n | 108 | 116 | 109 | 99 | 90 | 78 | 100 | 79 | 88 | 79 | 100 | 100 |
| Genotypes | AA: 0.55 | CC: 0.42 | CC: 0.36 | GG: 0.47 | TT:0.74 | GG: 0.67 | TT: 0.43 | CC: 0.42 | AA: 0.56 | TT: 0.91 | CC: 0.42 | CC: 0.43 |
| | GA: 0.45 | TC: 0.44 | CG: 0.49 | GT: 0.40 | TC:0.26 | GT: 0.30 | TG: 0.45 | CT: 0.47 | AG: 0.31 | TC: 0.05 | CA: 0.48 | CT: 0.41 |
| | GG: 0 | TT: 0.14 | GG: 0.15 | TT: 0.13 | CC: 0 | TT: 0.03 | GG: 0.12 | TT: 0.11 | GG: 0.13 | CC: 0.04 | AA: 0.10 | TT: 0.16 |
| Machado lineage, n | 43 | 49 | 48 | 40 | 35 | 29 | 39 | 31 | 34 | 28 | 34 | 39 |
| Genotypes | AA: 0.56 | CC: 0.76 | CC: 0.75 | GG: 0.83 | TT:0.69 | GG:0.66 | TT:0.77 | CC:0.77 | AA:0.82 | TT:1 | CC:0.85 | CC:0.79 |
| | GA: 0.44 | TC: 0.24 | CG: 0.25 | GT: 0.17 | TC:0.31 | GT:0.34 | TG:0.23 | CT:0.23 | AG:0.18 | TC:0 | CA:0.15 | CT:0.21 |
| | GG: 0 | TT: 0 | GG: 0 | TT: 0 | CC:0 | TT:0 | GG:0 | TT:0 | GG:0 | CC:0 | AA:0 | TT:0 |
| Joseph lineage, n | 41 | 43 | 38 | 36 | 33 | 30 | 36 | 30 | 32 | 31 | 36 | 36 |
| Genotypes | AA: 0.56 | CC: 0 | CC: 0 | GG: 0 | TT:0.73 | GG:0.83 | TT:0.25 | CC:0.23 | AA:0.25 | TT:0.90 | CC:0.22 | CC:0.28 |
| | GA: 0.44 | TC: 0.70 | CG: 0.71 | GT:0.72 | TC:0.27 | GT:0.17 | TG:0.75 | CT:0.77 | AG:0.53 | TC:0.10 | CA:0.78 | CT:0.72 |
| | GG: 0 | TT: 0.30 | GG: 0.29 | TT:0.28 | CC:0 | TT:0 | GG:0 | TT:0 | GG:0.22 | CC:0 | AA:0 | TT:0 |

**b.**

| Portuguese Mainland MJD cohort | | | |
|---|---|---|---|
| rs16999141 | rs1048755 | rs12895357 | rs7158733 |
| 108 | | | |
| AA: 0.48 | CC: 0.51 | CC: 0.49 | GG: 0.51 |
| GA: 0.52 | TC: 0.42 | CG: 0.44 | GT: 0.42 |
| GG: 0 | TT: 0.07 | GG: 0.07 | TT: 0.07 |
| 73 | | | |
| AA: 0.47 | CC: 0.75 | CC: 0.73 | GG: 0.75 |
| GA: 0.53 | TC: 0.25 | CG: 0.27 | GT: 0.25 |
| GG: 0 | TT: 0 | GG: 0 | TT: 0 |
| 35 | | | |
| AA: 0.51 | CC: 0 | CC: 0 | GG: 0 |
| GA: 0.49 | TC: 0.77 | CG: 0.77 | GT: 0.77 |
| GG: 0 | TT: 0.23 | GG: 0.23 | TT: 0.23 |

**c.**

| | rs16999141 | rs1048755 | rs12895357 | rs7158733 | rs11628764 | rs10151135 | rs3092822 | rs709930 | rs1055996 | rs55966267 | rs910369 | rs7158238 |
|---|---|---|---|---|---|---|---|---|---|---|---|---|
| | European population (1000 Genomes Project, Phase 3)* | | | | | | | | | | | |
| Genotypes n=503 | AA: 0.22 | CC: 0.62 | CC: 0.52 | GG: 0.62 | TT:0.49 | GG: 0.81 | TT: 0.62 | CC: 0.62 | AA: 0.62 | TT: 0.96 | CC: 0.62 | CC: NA |
| | GA: 0.46 | CT: 0.32 | CG: 0.41 | GT: 0.32 | TC:0.41 | GT: 0.18 | TG: 0.32 | CT: 0.32 | AG: 0.32 | TC: 0.04 | CA: 0.31 | CT: NA |
| | GG: 0.32 | TT: 0.07 | GG: 0.07 | TT: 0.07 | CC: 0.10 | TT: 0.01 | GG: 0.10 | TT: 0.10 | GG: 0.10 | CC: 0.00 | AA: 0.07 | TT: NA |
| Alleles n=1006 | A: 0.45 | C: 0.78 | C: 0.73 | G: 0.78 | T:0.70 | G: 0.90 | T: 0.78 | C: 0.78 | A: 0.78 | T: 0.98 | C: 0.78 | C: NA |
| | G: 0.55 | T: 0.22 | G: 0.27 | T: 0.22 | C: 0.30 | T: 0.10 | G: 0.22 | T: 0.22 | G: 0.22 | C: 0.02 | A: 0.22 | T: NA |
| | East Asian population (1000 Genomes Project, Phase 3)* | | | | | | | | | | | |
| Genotypes n=504 | AA: 0.33 | CC: 0.33 | CC: 0.34 | GG: 0.33 | TT:0.97 | GG: 1.00 | TT: 0.32 | CC: 0.47 | AA: 0.49 | TT: 0.82 | CC: 0.47 | CC: NA |
| | GA: 0.46 | CT: 0.47 | CG: 0.47 | GT: 0.47 | TC:0.03 | GT: 0.00 | TG: 0.47 | CT: 0.42 | AG: 0.41 | TC: 0.17 | CA: 0.42 | CT: NA |
| | GG: 0.21 | TT: 0.21 | GG: 0.19 | TT: 0.21 | CC: 0.00 | TT: 0.00 | GG: 0.21 | TT: 0.11 | GG: 0.10 | CC: 0.01 | AA: 0.11 | TT: NA |
| Alleles n=1008 | A: 0.56 | C: 0.56 | C: 0.58 | G: 0.56 | T:0.99 | G: 100 | T: 0.56 | C: 0.68 | A: 0.70 | T: 0.91 | C: 0.68 | C: NA |
| | G: 0.44 | T: 0.44 | G: 0.42 | T: 0.44 | C: 0.01 | T: 0.00 | G: 0.44 | T: 0.32 | G: 0.30 | C: 0.09 | A: 0.32 | T: NA |

* European population includes CEU (Utah residents with Northern and Western European ancestry), FIN (Finnish in Finland), GBR (British in England and Scotland), IBS (Iberian population in Spain), and TSI (Toscani in Italia) sub-populations. East Asian population includes CHB (Han Chinese in Bejing, China), JPT (Japanese in Tokyo, Japan), CHS (Southern Han Chinese), CDX (Chinese Dai in Xishuanagbanna, China), and KHV (Kinh in Ho Chi Minh City, Vietnam) sub-populations.
NA: not available

In the Azorean MJD cohort, several SNVs (rs910369, rs7158733, rs3092822, rs7158238, rs709930, rs1055996, rs12895357) are in linkage disequilibrium with rs1048755 (Table 4) and, thus, do not provide additional information to distinguish between the two *ATXN3* $(CAG)_n$ alleles. Moreover, rs16999141 and rs12895357 cannot be used to assess *ATXN3* RNA levels, as outlined before.

In summary, we propose a set of five SNVs – rs1048755, rs10151135, rs11628764, rs910369, rs55966267 – that, together, enabled allele-specific expression analysis in ~59% of MJD samples (Figure 2).

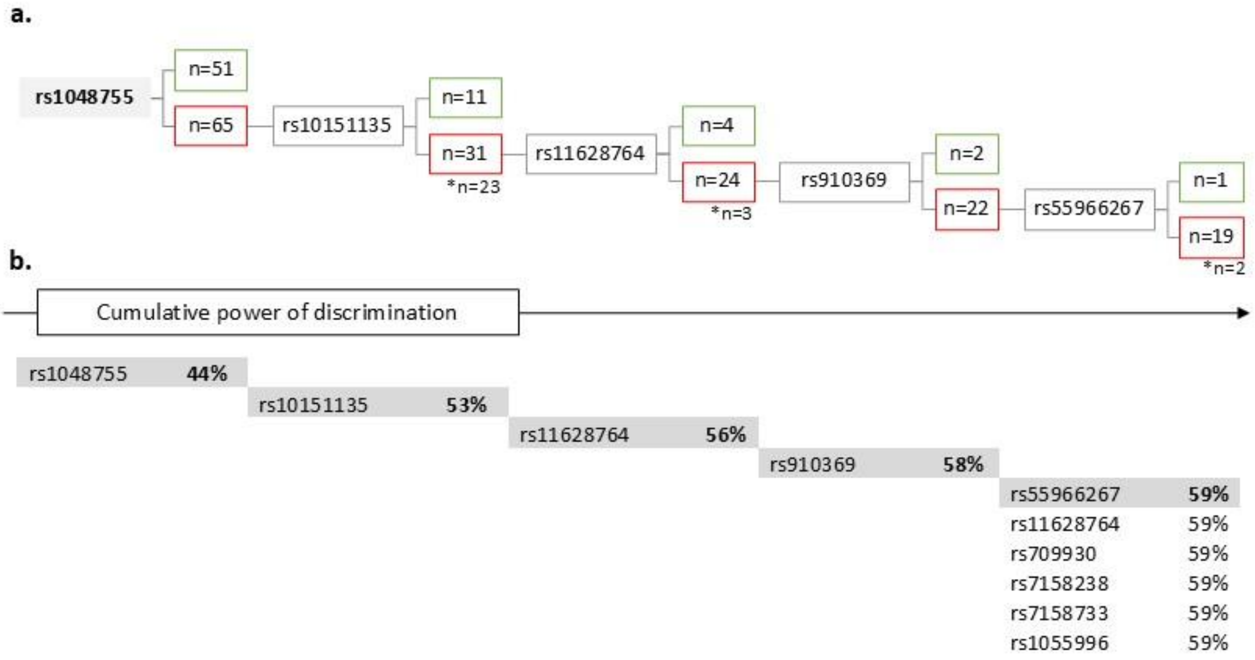


**Figure 2.** Discriminatory ability of five SNVs to differentiate normal and expanded RNA *ATXN3* alleles in MJD blood samples. (a) Decision tree illustrates the distribution of heterozygous and homozygous individuals for each SNV. Green box = number of heterozygous subjects; red box = number of homozygous subjects; * individuals lacking genotype information. (b) Cumulative discrimination power of SNVs (%) as additional variants are included to distinguish normal and expanded RNA *ATXN3* alleles.

**Table 4.** Genotypic linkage disequilibrium (LD) was assessed (a), and corresponding contingency tables were generated (b) for rs1048755 and the remaining 11 SNVs, comprising a total of 12 loci. LD was evaluated using the likelihood ratio test (G-test) under the null hypothesis (Ho) that genotypes at one locus are independent of genotypes at the other locus. Analyses were conducted using Arlequin version 4.7.5 (9,10).

**a.**

Genepop 4.7.5, Genotypic linkage disequilibrium

Number of populations detected: 1
Number of loci detected: 12

Markov chain parameters
Dememorisation: 1000
Batches: 100
Iterations per batch: 1000

| Locus#1 | Locus#2 | P-Value | S.E. | Switches |
|---|---|---|---|---|
| rs1048755 | rs910369 | 0 | 0 | 75700 |
| | rs7158733 | 0 | 0 | 77392 |
| | rs3092822 | 0 | 0 | 77017 |
| | rs7158238 | 0 | 0 | 77144 |
| | rs10151135 | 0.61961 | 0.00314 | 80184 |
| | rs709930 | 0 | 0 | 73496 |
| | rs55966267 | 0.33313 | 0.00240 | 61805 |
| | rs11628764 | 0.80657 | 0.00182 | 80443 |
| | rs1055996 | 0 | 0 | 74386 |
| | rs16999141 | 0.70031 | 0.00317 | 84451 |
| | rs12895357 | 0 | 0 | 77691 |

**b.**

Genepop 4.7.5, Contingency tables for genotypic disequilibrium

Number of populations detected: 1
Number of loci detected: 12

loci: rs1048755(G>A) and rs910369(C>A)

| | | rs1048755 | | |
|---|---|---|---|---|
| | | GA | AA | |
| rs910369 | CC | **1** | **37** | **38** |
| | CA | **42** | **7** | **49** |
| | AA | **0** | **9** | **9** |
| | | **43** | **53** | **96** |

loci: rs1048755(G>A) and rs10151135(G>T)

| | | rs1048755 | | |
|---|---|---|---|---|
| | | GA | AA | |
| rs10151135 | 1.1 | **23** | **29** | **52** |
| | 1.2 | **12** | **11** | **23** |
| | | **35** | **40** | **75** |

loci: rs1048755(G>A) and rs1055996(A>G)

| | | rs1048755 | | |
|---|---|---|---|---|
| | | GA | AA | |
| rs1055996 | 1.1 | **4** | **36** | **40** |
| | 1.2 | **27** | **0** | **27** |
| | 2.2 | **1** | **8** | **9** |
| | | **32** | **44** | **76** |

loci: rs1048755(G>A) and rs7158733(G>T)

| | | rs1048755 | | |
|---|---|---|---|---|
| | | GA | AA | |
| rs7158733 | 1.1 | **2** | **42** | **44** |
| | 1.2 | **40** | **0** | **40** |
| | 2.2 | **1** | **11** | **12** |
| | | **43** | **53** | **96** |

loci: rs1048755(G>A) and rs709930(C>T)

| | | rs1048755 | | |
|---|---|---|---|---|
| | | 1.2 | 2.2 | |
| rs709930 | 1.1 | **1** | **29** | **30** |
| | 1.2 | **32** | **5** | **37** |
| | 2.2 | **1** | **7** | **8** |
| | | **34** | **41** | **75** |

loci: rs1048755(G>A) and rs16999141(C>T)

| | | rs1048755 | | |
|---|---|---|---|---|
| | | GA | AA | |
| rs16999141 | 1.2 | **21** | **27** | **48** |
| | 2.2 | **28** | **30** | **58** |
| | | **49** | **57** | **106** |

loci: rs1048755(G>A) and rs3092822(T>G)

| | | rs1048755 | | |
|---|---|---|---|---|
| | | GA | AA | |
| rs3092822 | 1.1 | **3** | **40** | **43** |
| | 1.2 | **41** | **2** | **43** |
| | 2.2 | **0** | **11** | **11** |
| | | **44** | **53** | **97** |

loci: rs1048755(G>A) and rs55966267(T>C)

| | | rs1048755 | | |
|---|---|---|---|---|
| | | GA | AA | |
| rs55966267 | 1.1 | **32** | **39** | **71** |
| | 1.2 | **3** | **1** | **4** |
| | | **35** | **40** | **75** |

loci: rs12895357(C>G) and rs1048755(G>A)

| | | rs1048755 | | |
|---|---|---|---|---|
| | | GA | AA | |
| rs12895357 | 1.1 | **0** | **13** | **13** |
| | 1.2 | **43** | **11** | **54** |
| | 2.2 | **0** | **36** | **36** |
| | | **43** | **60** | **103** |

loci: rs1048755(G>A) and rs7158238(C>T)

| | | rs1048755 | | |
|---|---|---|---|---|
| | | GA | AA | |
| rs7158238 | 1.1 | **1** | **40** | **41** |
| | 1.2 | **40** | **2** | **42** |
| | 2.2 | **1** | **11** | **12** |
| | | **42** | **53** | **95** |

loci: rs1048755(G>A) and rs11628764(T>C)

| | | rs1048755 | | |
|---|---|---|---|---|
| | | GA | AA | |
| rs11628764 | 1.1 | **29** | **36** | **65** |
| | 1.2 | **11** | **11** | **22** |
| | | **40** | **47** | **87** |

Step 2: SNV-$(CAG)_n$ phasing

A subset of 24 cDNA samples from heterozygotes for rs1048755 (previously identified in Step 1) were used. Amplification of the *ATXN3*_$(CAG)_n$ fragment along with rs1048755 was performed by PCR using a novel set of primers, namely ATXN3-F (5'-AGTCCAGAGTATCAGAGGCTCA-3') and MJD-R (5'-AAGTGCTCCTGAACTGGTGG-3'), in a solution containing 1X MyFi Reaction Buffer (Meridian Bioscience), 0.2 uM of each primer, 2U of MyFi DNA Polymerase (Meridian Bioscience) and 100 ng of cDNA. Amplification started with an initial denaturation for 1 min at 95°C, followed by 30 sec at 95°C, 45 sec at 57°C and 1 min at 72°C for 39 cycles, and a final elongation step of 10 min at 72°C. The size of the PCR products was determined by agarose gel (2%) electrophoresis, in parallel with the GeneRuler 100 bp Plus DNA Ladder (ThermoFisher Scientific). Such PCR fragments (two per sample, corresponding to the non-expanded and expanded alleles) were then excised from TBE-buffered agarose gels and purified using the Zymoclean Gel DNA Recovery Kit (Zymo Research), following manufacturers' instructions. The sequence of nucleotides corresponding to the eluted fragments regarding non-expanded and expanded $(CAG)_n$ alleles along with the respective rs1048755 allele was assessed, by Sanger sequencing at Eurofins Genomics.

<u>Calculation of the relative expression ratio of expanded to non-expanded *ATXN3* alleles:</u> for samples from heterozygotes (genotype G/A), the relative expression of the two alleles was calculated as delta Ct (ΔCt) = Ct (expanded allele) − Ct (non-expanded allele). The ratio of the two alleles was estimated as $2^{-\Delta Ct}$.

REFERENCES

1. Dyer SC, Austine-Orimoloye O, Azov AG, et al. Ensembl 2025. Nucleic Acids Research 2025;53(D1):D948–D957. https://doi.org/10.1093/nar/gkae1071

2. Raposo M, Hübener-Schmid J, Tagett R, et al. Blood and cerebellar abundance of ATXN3 splice variants in spinocerebellar ataxia type 3/Machado-Joseph disease. Neurobiology of disease 2024;193:106456. https://doi.org/10.1016/j.nbd.2024.106456

3. Harrison PW, Amode MR, Austine-Orimoloye O, et al. Ensembl 2024. Nucleic acids research 2024;52(D1):D891–D899. https://doi.org/10.1093/nar/gkad1049

4. Sidky AM, Melo ARV, Kay TT, et al. Age-dependent somatic expansion of the ATXN3 CAG repeat in the blood and buccal swab DNA of individuals with spinocerebellar ataxia type 3/Machado-Joseph disease. Human genetics. https://doi.org/10.1007/s00439-024-02698-7

5. Melo ARV, Raposo M, Ventura M, et al. Genetic Variation in ATXN3 ( Ataxin - 3 ) 3 ' UTR : Insights into the Downstream Regulatory Elements of the Causative Gene of Machado - Joseph Disease / Spinocerebellar Ataxia Type 3. The Cerebellum;3(0123456789). https://doi.org/10.1007/s12311-021-01358-0

6. Costa IPD, Almeida BC, Sequeiros J, Amorim A, Martins S. A pipeline to assess disease-associated haplotypes in repeat expansion disorders: The example of MJD/SCA3 locus. Frontiers in Genetics 2019;10(FEB):1–9. https://doi.org/10.3389/fgene.2019.00038

7. Martins S, Calafell F, Gaspar C, et al. Asian origin for the worldwide-spread mutational event in Machado-Joseph disease. Archives of neurology 2007;64(10):1502–1508. https://doi.org/10.1001/archneur.64.10.1502

8. Excoffier L, Lischer HEL. Arlequin suite ver 3.5: a new series of programs to perform population genetics analyses under Linux and Windows. Molecular ecology resources 2010;10(3):564–567. https://doi.org/10.1111/j.1755-0998.2010.02847.x

9. Raymond M, Rousset F. AN EXACT TEST FOR POPULATION DIFFERENTIATION. Evolution; international journal of organic evolution 1995;49(6):1280–1283. https://doi.org/10.1111/j.1558-5646.1995.tb04456.x

10. Rousset F. genepop'007: a complete re-implementation of the genepop software for Windows and Linux. Molecular ecology resources 2008;8(1):103–106. https://doi.org/10.1111/j.1471-8286.2007.01931.x

**Supplementary Figure 1.** Characterization of the Azorean MJD cohort by sex (a), age at evaluation (b), $(CAG)_n$ repeat size in the expanded allele (c), and age at onset (d). The age at onset among MJD lineages was estimated using ANCOVA, with the $(CAG)_n$ repeat length of the expanded allele (72 repeats) included as a covariate (e). Data in panels (b), (c) and (d) are presented as volcano plots, while data in panel (e) are shown as mean ± standard error of mean.

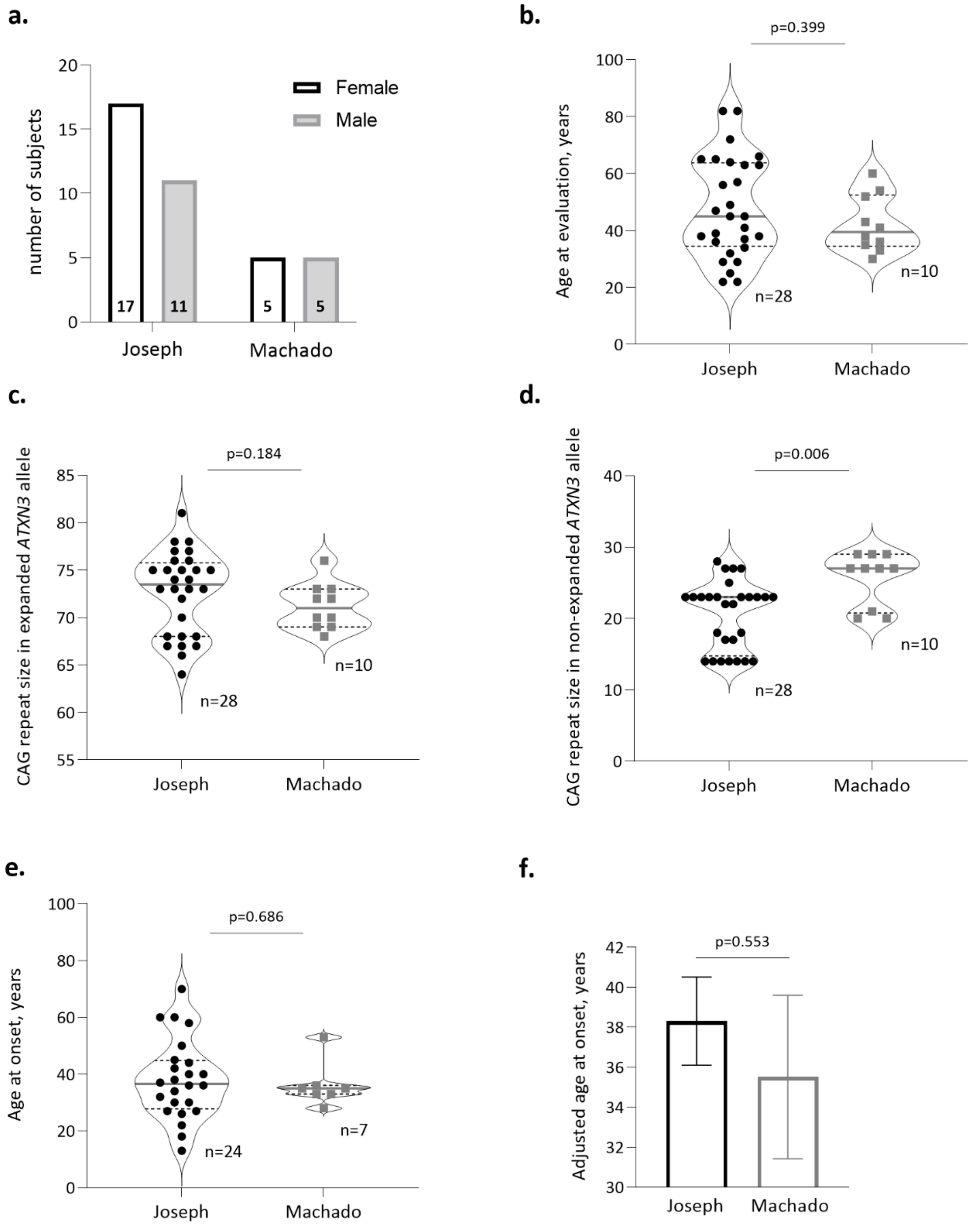
